# Composite endpoints to detect treatment effects on MS disability progression. Lessons from phase III trial data

**DOI:** 10.64898/2026.04.22.26351458

**Authors:** Francesca Bovis, Noemi Montobbio, Alessio Signori, Tomas Kalincik, Douglas L Arnold, Mar Tintorè, Ludwig Kappos, Maria Pia Sormani

## Abstract

Disability worsening is the critical long-term outcome in multiple sclerosis, yet the Expanded Disability Status Scale incompletely captures neurological deterioration and has limited sensitivity in the short time windows of clinical trials. Composite endpoints incorporating functional measures have been proposed to address these limitations, but whether they reliably improve detection of treatment effects has not been established across trials.

We conducted a post-hoc analysis of individual patient data from ten phase III randomised controlled trials (ASCEND, BRAVO, CONFIRM, DEFINE, EXPAND, INFORMS, OLYMPUS, OPERA I/II, and ORATORIO; n = 9,369), spanning relapsing-remitting and progressive multiple sclerosis. Confirmed disability worsening was defined using harmonised criteria with the msprog package and confirmed at 24 weeks. Treatment effects were estimated using Cox proportional hazards models and combined across trials in a one-stage individual patient data framework. Composite endpoints were constructed from the Expanded Disability Status Scale, the timed 25-foot walk test, and the nine-hole peg test using logical unions (OR-type), intersections (AND-type), and majority-vote structures. Sensitivity to treatment effect was quantified using Z-scores (the ratio of the pooled log-hazard ratio to its standard error) and compared to the Expanded Disability Status Scale reference using interaction tests.

Event rates varied across components: the timed walk test generated the highest rates (up to 46.8%) while the nine-hole peg test generated the lowest (as low as 2.1%). OR-type composite endpoints showed weaker treatment effects than the Expanded Disability Status Scale alone, with the largest reductions in sensitivity observed for endpoints incorporating the timed walk test (ΔZ up to +2.26; interaction p = 0.004). These findings were confirmed across disease subtypes and were pronounced in relapsing-remitting trials, where no composite endpoint outperformed the Expanded Disability Status Scale. In progressive multiple sclerosis, the combination of the Expanded Disability Status Scale and the nine-hole peg test showed numerically stronger treatment effects (ΔZ = −1.65), though interaction tests did not reach statistical significance (p = 0.051).

Composite endpoints do not systematically improve treatment effect detection in multiple sclerosis trials. Increased event capture driven by the timed walk test introduces noise that dilutes the treatment signal rather than amplifying it, highlighting that event rate and endpoint quality are not interchangeable. Upper limb function assessed by the nine-hole peg test provides complementary and specific information, particularly in progressive disease. The combination of global disability and upper limb measures represents a promising direction for future endpoint development in progressive multiple sclerosis trials, warranting validation.

## Introduction

Multiple sclerosis (MS) is a chronic and often disabling neurological condition characterized by a progressive loss of neurological function. The long-term clinical outcome of MS is primarily determined by the extent of disability progression rather than the frequency of relapses.^1^ While several very effective disease-modifying treatments (DMTs) have been approved to reduce relapse rates, the more pressing and unmet clinical need is to find more effective ways to limit the progression of disability. This shift in focus has made the accurate assessment of disability worsening a critical endpoint in MS clinical trials.

The Expanded Disability Status Scale (EDSS) has traditionally been the primary tool used to evaluate disability worsening in MS trials. Despite the availability and widespread use of a better standardized version (Neurostatus-EDSS) that allows for more consistent assessments and higher reproducibility between different raters, the EDSS has notable limitations. In particular, it does not fully capture the entire spectrum of disability progression and has low sensitivity for detecting subtle changes over time.^2-6^ These limitations are particularly relevant over short-term periods of 2 to 3 years, the typical duration for clinical trials.

Furthermore, because EDSS-defined disability progression events are relatively rare, occurring in only about 10% of patients over two years in the relapsing–remitting phase, large sample sizes are required to achieve adequate statistical power. In MS, two identically designed phase 3 trials are typically conducted for regulatory purposes: each trial is powered for relapse rates, while the disability endpoint is assessed in the pooled analysis of both studies. This requirement poses significant logistical and financial challenges, which have become even more relevant as the use of placebo is no longer considered ethical, making active-controlled arms necessary.

To address these shortcomings, alternative functional performance measures such as the timed 25-foot walk test (T25FWT) and the 9-hole peg test (9HPT) have been incorporated in a composite definition of progression in recent MS clinical trials.^7-9^ The T25FWT evaluates walking ability and walking speed over a standardized distance, while the 9HPT assesses fine motor skills, specifically coordination, and cognitive-motor integration of the upper extremities. Since these tests measure distinct aspects of neurological function, they only partially overlap and may complement the worsening detected by EDSS. This suggests that combining multiple such measures into composite endpoints could provide a more sensitive and reliable assessment of overall disability worsening in MS. The general feeling is that composite endpoints have the potential to improve precision in the estimated treatment effect, increase sensitivity to change, and lower the sample size requirements needed to detect significant treatment effects, but there is conflicting evidence about that.

In 2014, Zhang et al.^10^ evaluated in a post-hoc analysis of the OLYMPUS trial,^11^ a study evaluating rituximab vs placebo in primary progressive MS, the performance of composite endpoints that integrate results from EDSS, T25FWT, and 9HPT to enhance the detection of treatment effect. By combining multiple measures, some of these composite endpoints were shown to increase the sensitivity to detect the treatment effect on disability worsening, with higher treatment effect estimates and smaller p-value.^10^ More recently, Kappos et al.^12^ systematically evaluated the predictive validity of composite confirmed disability worsening, demonstrating that early worsening in performance measures like T25FWT and 9HPT strongly predicts subsequent EDSS progression, thus supporting the clinical utility of adding performance measures as components to disability outcomes in MS trials.

The goal of the present study is to determine whether one or more composite outcomes are systematically more sensitive than EDSS alone in estimating treatment effects on confirmed disability worsening (CDW) in MS trials. The analysis was conceived within a clinical trial framework, under the assumption of a true therapeutic effect, to identify the combination of endpoints most sensitive to capturing such benefit.

We conducted a post-hoc analysis of 10 phase III MS trials evaluating the efficacy of different DMTs on CDW across various MS subtypes. We evaluated composite outcomes previously proposed in the literature and additionally constructed alternative composite definitions to explore whether different combinations of endpoints could further enhance sensitivity to treatment effects. Beyond traditional physical disability measures, we incorporated a cognitive assessment into the composite endpoints to provide a more comprehensive evaluation of disability worsening.

## Materials and methods

### Data

Data were pooled from ten randomized clinical trials: ASCEND (ClinicalTrials.gov identifier numbers: NCT01416181), BRAVO (NCT00605215), CONFIRM (NCT00451451), DEFINE (NCT00420212), EXPAND (NCT01665144), INFORMS (NCT00731692), OLYMPUS (NCT00087529), OPERA I (NCT01247324), OPERA II (NCT01412333), and ORATORIO (NCT01194570).^11,13-20^ OPERA I/II, DEFINE, CONFIRM and BRAVO were trials in RRMS that compared different treatments: OPERA I/II evaluated ocrelizumab versus subcutaneous interferon beta-1a (IFNβ-1a); DEFINE and CONFIRM assessed dimethyl fumarate against both glatiramer acetate and placebo or placebo alone; and BRAVO tested laquinimod against both intramuscular IFNβ-1a and placebo.

ORATORIO, OLYMPUS, and INFORMS were placebo-controlled trials in PPMS, investigating the efficacy of ocrelizumab, rituximab, and fingolimod, respectively. Lastly, ASCEND and EXPAND were placebo-controlled trials in SPMS, examining the effects of natalizumab and siponimod, respectively.

### Study outcomes

Worsening on the EDSS was defined as an EDSS increase of 1.5 points if baseline=0, 1 if 0<baseline<=5.0, 0.5 if baseline>5.0 and worsening on the 9HPT and on the T25FW was defined as an increase of 20% of the baseline value. Cognitive decline was defined as either a 4-point decline or 20% worsening from the reference values for Symbol Digit Modalities Test (SDMT) or Paced Auditory Serial Addition Test (PASAT) if SDMT values were unavailable. In the primary analysis, all the outcomes were confirmed at 24 weeks. A sensitivity analysis was run for 12-week CDW (Supplementary Material). The study outcomes, specifically 24-week CDW measured by different combinations of EDSS, T25FW, and 9HPT (based on the mean of both hands), as detailed below, were recalculated for each trial using a common definition of CDW, harmonized according to the default parameters of the “msprog” package (Appendix in Supplementary Material). This ensured consistent derivation of confirmed worsening for EDSS and all composite endpoints.^21^

The time to CDW was defined as the period from randomization to the initial occurrence of CDW. If a patient had not experienced CDW (including confirmation) by their final scheduled visit, the time to CDW was censored at the date of the last available assessment.

### Composite outcomes

Composite endpoints were constructed from the primary CDW components (EDSS, T25FWT, and 9HPT) using logical unions (∪, corresponding to “OR”) and intersections (∩, corresponding to “AND”).

If multiple CDW events occurred across the components of a composite endpoint, the composite event time was defined according to the prespecified logical structure. For OR-type composites, the event time corresponded to the earliest occurrence of any component-specific CDW. For AND-type composites, the event time was defined as the latest occurrence among the required component-specific CDWs (i.e., the time at which all criteria were satisfied). For k-of-n composites, the event time was defined as the time at which the kth component-specific CDW was confirmed.

CDW rates, treatment effects (expressed as Hazard Ratios (HR)), Z-values, and p-values were estimated for a set of predefined composite endpoints. These included composite definitions previously described in the literature and used in clinical trials so far (1), as well as additional configurations constructed to systematically vary the logical combination of components (2) and the inclusion of cognitive assessment (3).

#### 1. Primary analysis: endpoints and their OR-type composite definitions

The primary analysis was run on the 24-week CDW of the following endpoints (previously used as endpoints in MS clinical trials):

- **EDSS**
- **T25FWT**
- **9HPT**
- **EDSS** ∪ **T25FWT** ∪ **9HPT** (CDW from at least one of the three tests)
- **EDSS** ∪ **T25FWT** (CDW from either EDSS or T25FWT)
- **EDSS** ∪ **9HPT** (CDW from either EDSS or 9HPT)

#### 2. Exploratory analysis: AND-type composite definition

An exploratory analysis was run on the 24-week CDW of the following endpoints that can be explored as potential outcomes in future clinical trials:

- **EDSS** ∩ **9HPT** requires confirmed worsening in both EDSS and 9HPT
- **EDSS** ∩**T25FWT** requires confirmed worsening in both EDSS and T25FWT
- **EDSS** ∩ **(T25FWT** ∪ **9HPT)** requiring EDSS worsening as a mandatory component
- ≥**2 of (EDSS, T25FWT, 9HPT)** requiring worsening in at least two of the three assessed measures.
- **EDSS** ∩ **T25FWT** ∩ **9HPT**, with CDW defined by worsening in all three tests.

#### 3. Exploratory analysis: Expanded composites including cognitive assessment

Finally, an exploratory analysis was conducted on 24-week CDW across all previously defined endpoints, also incorporating worsening in a cognitive outcome (COG) as a potential additional endpoint for future clinical trials:

- **EDSS** ∪ **T25FWT** ∪ **9HPT** ∪ COG (CDW from any of the three tests or cognitive decline).
- **EDSS** ∪ **T25FWT** ∪ **COG** (CDW from either EDSS, T25FWT, or cognitive decline).
- **EDSS** ∪ **9HPT** ∪ **COG** (CDW from either EDSS, 9HPT, or cognitive decline).
- **EDSS** ∩ **9HPT** ∩ **COG** (CDW in EDSS and 9HPT and cognitive decline).
- **EDSS** ∩ **T25FWT** ∩ **COG** (CDW in EDSS and T25FWT and cognitive decline).
- **(EDSS** ∩ **T25FWT)** ∪ **(EDSS** ∩ **9HPT)** ∪ **(EDSS** ∩ **COG)** (CDW in EDSS confirmed by a CDW in either T25FWT, 9HPT, or cognitive decline).
- **(EDSS** ∩ **T25FWT)** ∪ **(EDSS** ∩ **9HPT)** ∪ **(T25FWT** ∩ **9HPT)** ∪ **(EDSS** ∩ **COG)** ∪ **(T25FWT** ∩ **COG)** ∪ **(9HPT** ∩ **COG)** (CDW confirmed by any two of the tests, including cognitive decline).
- **EDSS** ∩ **T25FWT** ∩ **9HPT** ∩ **COG** (CDW confirmed by all three tests and cognitive decline).

### Statistical analysis

For each trial and endpoint, treatment effects were estimated using Cox proportional hazards models applied to time to CDW, defined according to each specific endpoint definition, based on individual patient data from all included clinical trials. Treatment effects were summarized as hazard ratios (HRs) with their corresponding standard errors (SE) and 95% confidence intervals (CI).

To compare treatment effects across endpoint definitions, individual patient data from all trials were analyzed jointly using Cox proportional hazards models stratified by trial and outcome type to allow for trial-specific baseline hazards. In this one-stage approach, treatment effects were estimated within each trial and combined within a single model, with trials contributing proportionally to their information content (i.e., number of observed events) through the partial likelihood.

Robust variance estimation was obtained by clustering on patient identifiers. The model included an interaction term between treatment arm and endpoint definition to assess whether the estimated treatment effect differed when CDW was defined using a composite endpoint compared with EDSS alone. To summarize the performance of each composite endpoint as compared to EDSS alone across trials, mixed-effects models were fitted to the log(HR) values using inverse-variance weighting (weights = 1/SE^2^), with trial included as a random intercept. To rank the overall performance, Z-scores were calculated as the ratio of the pooled log(HR) to its standard error. The Z-score incorporates both the magnitude of the treatment effect (expressed as the log HR) and its variability (SE) thereby reflecting the balance between signal (effect size) and noise (statistical uncertainty) for each endpoint.

Pre-specified subgroup analyses were performed separately for relapsing-remitting MS (RRMS) and progressive MS trials. The same statistical approach was applied in sensitivity analyses applied to 12-week CDW and in exploratory analyses examining different composite endpoints (Supplementary Material).

### Data availability

Individual patient data from these RCTs were made available through the International Progressive MS Alliance (IPMSA) project (award reference number PA-1603-08175). Access requests should be forwarded to the relevant data controllers.

## Results

The baseline demographic and clinical characteristics of participants in each RCT were representative of the MS populations targeted in these studies. Detailed descriptions of these characteristics can be found in the respective published articles and are not included here.^11,13-20^ The treatment effect on 24-week CDW reported in the trial publications is summarized in the Supplementary Table 1 in the Appendix.

### Confirmed Disability Worsening events

The number of CDW events for each endpoint definition across the trials is reported in Table 1. Overall, the T25FWT showed the highest number of events (with event rates ranging from 9.09% to 46.82%), whereas the lowest rates were observed for the 9HPT (ranging from 2.06% to 19.92%).

Table 2 presents the relative contribution of each component to CDW within the pooled trial cohorts. The T25FWT was the primary driver of progression in composite endpoints that included it: 49.6% of CDW events based on EDSS ∪ T25FWT ∪ 9HPT were attributable to T25FWT events, while 38.3% were driven by EDSS and only 12.1% by 9HPT. T25FWT accounted for 56.5% of events in the EDSS ∪ T25FWT composite, whereas EDSS contributed 76.6% of events in the EDSS ∪ 9HPT composite. Overall, the relative contribution of individual components was consistent across different confirmation window durations (see Supplementary Table 2).

### Treatment effects assessed by composite endpoints vs EDSS-based CDW

#### 1. OR-type composite definitions (Primary analysis)

Table 3 (and Supplementary Table 3) reports the ability of each composite endpoint to detect the treatment effect relative to CDW based on EDSS alone. The Z-score obtained for EDSS-based CDW was used as the reference, and results are expressed as differences in Z-score (ΔZ) of each composite endpoint compared with Z-score of EDSS alone. More negative Z-scores indicate a higher treatment effect–to–noise ratio, reflecting a greater benefit of the treatment arm compared with the control arm. Accordingly, negative ΔZ values indicate improved performance relative to EDSS, whereas positive ΔZ values indicate reduced sensitivity in detecting the treatment effect.

Across individual trials, results were heterogeneous. In three of the nine trials (BRAVO, DEFINE, and EXPAND), none of the alternative CDW definitions outperformed EDSS, as all endpoints showed positive ΔZ values. In four trials (ASCEND, INFORMS, ORATORIO, and CONFIRM), the largest improvements relative to EDSS (i.e., most negative ΔZ values) were observed for CDW defined using 9HPT alone. The OLYMPUS trial was the only study in which the greatest improvement relative to EDSS was observed for CDW defined using the T25FWT alone. In the OPERA trial, the largest improvement relative to EDSS was observed for the EDSS ∪ 9HPT composite endpoint.

Fig. 1 (and Supplementary Table 4) summarizes the integrated analysis across all trials, with p-values derived from the individual-patient interaction analysis.

**Figure 1.**
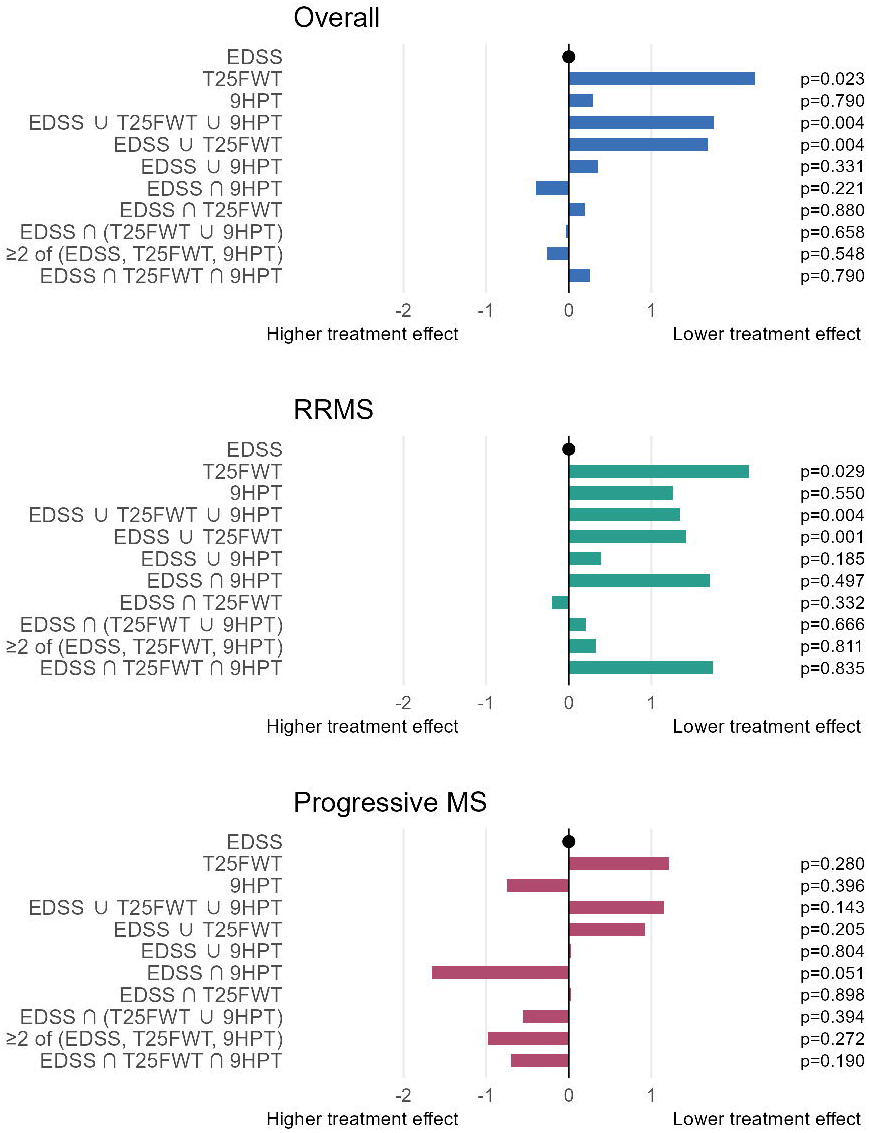
Differences in treatment effect estimates (ΔZ-scores relative to EDSS) for single and composite CDW endpoints in the overall population (A), relapsing remitting (B), and progressive MS (C). P-values represent interaction tests versus EDSS.

In the overall population, most endpoints showed positive ΔZ values, indicating weaker treatment effects compared with EDSS. The largest reductions in sensitivity were observed for T25FWT (ΔZ = +2.26) and for composites including T25FWT (e.g., ΔZ = +1.76 for EDSS ∪ T25FWT ∪ 9HPT and ΔZ = +1.68 for EDSS ∪ T25FWT). In contrast, 9HPT alone (ΔZ = +0.30) and the EDSS ∪ 9HPT composite (ΔZ = +0.35) showed values closer to zero, indicating performance more comparable to EDSS (Fig 1A). Interaction tests confirmed significantly weaker treatment effects for endpoints incorporating T25FWT compared with EDSS (p = 0.023 for T25FWT; p = 0.004 for EDSS ∪ T25FWT ∪ 9HPT and for EDSS ∪ T25FWT).

In analyses stratified by disease phenotype, similar patterns were observed. In RRMS, all composite endpoints showed positive ΔZ values relative to EDSS, indicating consistently weaker treatment effects, particularly for those including T25FWT (e.g., ΔZ = +2.18 for T25FWT and ΔZ = +1.42 for EDSS ∪ T25FWT; interaction p-values = 0.029) (Fig. 1B). In progressive MS, 9HPT alone (ΔZ = −0.75) and the EDSS ∪ 9HPT composite (ΔZ = +0.03) showed values close to zero, suggesting treatment effects broadly comparable to EDSS, whereas endpoints incorporating T25FWT again showed higher ΔZ values (e.g., ΔZ = +1.21), indicating weaker effects (Fig. 1C).

#### 2. AND-type composite definitions (Exploratory analysis)

In the overall population, several AND-type composite endpoints showed negative ΔZ values, indicating stronger treatment effects relative to EDSS. The largest improvements were observed for EDSS ∩ 9HPT (ΔZ = −0.40) and for the ≥2 of (EDSS, T25FWT, 9HPT) composite (ΔZ = −0.26), followed by EDSS ∩ (T25FWT ∪ 9HPT) (ΔZ = −0.04). In contrast, EDSS ∩ T25FWT (ΔZ = +0.19) and the stricter EDSS ∩ T25FWT ∩ 9HPT composite (ΔZ = +0.26) showed values close to or above zero, indicating little or no improvement over EDSS. However, interaction tests did not indicate statistically significant differences between EDSS and any composite endpoint (all interaction p-values > 0.05) (Figure 1A and Supplementary Table 4).

In RRMS, most composite endpoints showed ΔZ values close to or above zero, indicating no improvement or slightly weaker effects compared with EDSS. EDSS ∩ T25FWT showed a small negative ΔZ (−0.20), suggesting a modest numerical improvement, although not statistically significant (Figure 1B and Supplementary Table 4).

In progressive MS, several composite endpoints showed negative ΔZ values, indicating stronger treatment effects relative to EDSS. The largest improvements were observed for EDSS ∩ 9HPT (ΔZ = −1.65) and for the ≥2 composite (ΔZ = −0.98), while other combinations showed more modest differences (e.g., ΔZ = −0.55 for EDSS ∩ (T25FWT ∪ 9HPT)). Despite these numerical trends, interaction tests did not demonstrate statistically significant effect modification (all interaction p-values > 0.05), although a borderline interaction was observed for EDSS ∩ 9HPT (p = 0.051) (Figure 1C and Supplementary Table 4).

Results for expanded composite endpoints incorporating cognitive assessment are reported in Supplementary Table 5.

## Discussion

Disability progression remains a challenging endpoint in MS clinical trials. Although CDW event based on EDSS has long been the standard primary endpoint, its limited sensitivity and incomplete coverage of disability domains have motivated the development of composite endpoints incorporating functional performance measures such as the T25FWT and the 9HPT. In this analysis of ten phase III trials across relapsing-remitting and progressive MS, we systematically evaluated whether these composite definitions improve the detection of treatment effects compared with EDSS alone.

A key finding is the marked heterogeneity across individual components in both event rates and their contribution to composite outcomes. The T25FWT consistently generated the highest number of events, whereas the 9HPT showed substantially lower rates. However, a higher event frequency did not translate into improved sensitivity to treatment effects. In fact, endpoints driven by T25FWT systematically showed weaker treatment effects compared with EDSS. This suggests that the T25FWT, while highly responsive, may capture short-term fluctuations and non-specific variability, thereby introducing noise and diluting the treatment signal.

Consistently, we did not observe a systematic advantage of standard OR-type composite endpoints over EDSS. Although these composites increased the number of detected CDW events, this was largely driven by the T25FWT component and did not improve statistical power. These findings highlight an important methodological point: increasing event rates alone is not sufficient to enhance the ability to detect treatment effects. The quality and specificity of the events captured are equally, if not more, important.

This limitation may be further amplified by the dichotomization of continuous measures (e.g., ≥20% worsening), which can increase variability and reduce statistical efficiency. Alternative approaches based on continuous modeling or clinically meaningful thresholds may provide a more efficient use of these measures, although their applicability in regulatory settings remains uncertain.

When stratified by disease phenotype, distinct patterns emerged. In RRMS, EDSS alone consistently provided the strongest signal, outperforming all composite definitions. This supports its continued use as the primary disability endpoint in this population.

In contrast, in progressive MS, the 9HPT showed a treatment effect signal comparable to or stronger than EDSS. This finding was consistent across analyses and aligns with the evolving understanding of progressive disease, where upper limb dysfunction becomes more prominent and less tightly correlated with ambulation. These results reinforce the clinical relevance of the 9HPT and its potential to capture aspects of disability progression not adequately reflected by EDSS.

However, the broader use of the 9HPT is currently limited by the lack of standardization across trials, including differences in hand selection and outcome definition. Harmonization of 9HPT acquisition and analysis is therefore essential to ensure comparability and reproducibility in future studies.

We also explored alternative composite structures, including AND-type definitions and composites incorporating cognitive measures. AND-type composites showed promising signals, particularly in progressive MS, where requiring concordant worsening across domains may better reflect true, clinically meaningful progression. In this context, the combination of EDSS and 9HPT emerged as a particularly informative endpoint, showing a numerically stronger treatment effect compared with EDSS alone. This suggests that integrating global disability and upper limb function may enhance the detection of treatment effects in progressive disease.

More broadly, these findings highlight the importance of balancing sensitivity and specificity when designing composite endpoints. OR-type composites maximize sensitivity but may include less specific events, whereas AND-type composites improve specificity at the cost of fewer events. The optimal endpoint is therefore not the one with the highest event rate, but the one that maximizes the signal-to-noise ratio.

Overall, our results indicate that currently used composite endpoints do not systematically improve treatment effect detection compared with EDSS alone, particularly when driven by T25FWT. In contrast, specific components such as the 9HPT provide meaningful complementary information, especially in progressive MS.

Importantly, our analysis highlights a promising role for the composite endpoint based on EDSS and 9HPT in progressive MS trials. This combination appears to better capture clinically relevant multidomain worsening and may enhance sensitivity to treatment effects compared with EDSS alone.

These findings have important implications for future clinical trial design. In particular, they support the need to incorporate the EDSS and 9HPT endpoint in upcoming trials, both to validate these results and to provide more appropriate outcome measures for detecting treatment effects in progressive MS.

## Supplementary material

Supplementary material is available at *Brain* online.

