## Supplementary material for "Composite endpoints to detect treatment effects on MS disability progression. Lessons from phase III trial data"

**Appendix**

**Outcome definition according to the default parameters of “msprog” package:**

MSprog (

Data= data.frame containing longitudinal data (including: subject ID, outcome value, date of visit),

subj_col= Name of data column with subject ID,

value_col= Name of data column with outcome value,

date_col= Name of data column with date of visit,

outcome = Specifies the outcome type ('edss'; 'nhpt'; 't25fw'; 'sdmt'),

relapse = data.frame containing longitudinal data (including: subject ID and relapse date),

rsubj_col = Name of subject ID column for relapse data, if different from outcome data,

rdate_col = Name of onset date column for relapse data, if different from outcome data,

renddate_col = NULL,

subjects = NULL,

delta_fun = Minimum clinically meaningful change from the provided baseline value. Specifically:

• EDSS: 1.5 if baseline=0, 1 if 0<baseline<=5.0, 0.5 if baseline>5.0;

• NHPT and T25FW: 20% of baseline;

• SDMT: either 4 points or 20% of baseline,

worsening = NULL,

event = "firstCDW",

baseline = "fixed",

proceed_from = "firstconf",

sub_threshold_rebl = "none",

bl_geq = F,

relapse_rebl = F,

skip_local_extrema = "none",

validconf_col = NULL,

conf_days = 12 * 7,

conf_tol_days = c(7, 2 * 365.25),

require_sust_days = 0,

check_intermediate = T,

relapse_to_bl = 30,

relapse_to_event = 0,

relapse_to_conf = 30,

relapse_assoc = 90,

relapse_indep = NULL,

impute_last_visit = 0,

date_format = NULL,

include_dates = F,

include_value = F,

include_stable = T,

verbose = 1

)

**Supplementary Table 1.** **Published hazard ratios and 95% confidence intervals for confirmed disability worsening and functional outcomes at 12- and 24-week confirmation across clinical trials**

| **Trial** | **CDW** | **N** | **N (treatment arm)** | **N (placebo arm)** | **HR (95%CI) published** |
| --- | --- | --- | --- | --- | --- |
| **ASCEND** | EDSS 12-week | 889 | 440 | 449 | 1.06 (0.74-1.53)* |
| **ASCEND** | T25FW 12-week | 889 | 440 | 449 | 0.98 (0.74-1.30)* |
| **ASCEND** | 9HPT 12-week | 889 | 440 | 449 | 0.56 (0.40-0.80)* |
| **BRAVO** | EDSS 12-week | 884 | 434 | 450 | 0.69 (0.46-1.02) |
| **BRAVO** | EDSS 24-week | 884 | 434 | 450 | 0.61 (0.38-0.98)* |
| **CONFIRM** | EDSS 12-week | 1070 | 707 | 363 | 0.79/0.76 p=0.2 |
| **DEFINE** | EDSS 12-week | 1234 | 826 | 408 | 0.62/0.66 p=0.01 |
| **EXPAND** | EDSS 12-week | 1645 | 1099 | 546 | 0.79 (0.65-0.95) |
| **EXPAND** | EDSS 24-week | 1645 | 1099 | 546 | 0.74 (0.60-0.92) |
| **EXPAND** | T25FW 12-week | 1645 | 1099 | 546 | 0.94 (0.80-1.10) |
| **INFORMS** | EDSS 12-week | 970 | 483 | 487 | 0.88 (0.72-1.08) |
| **INFORMS** | 9HPT 12-week | 970 | 483 | 487 | 0.93 (0.71-1.22) |
| **INFORMS** | T25FW 12-week | 970 | 483 | 487 | 0.94 (0.78-1.14) |
| **OLYMPUS** | EDSS 12-week | 439 | 292 | 147 | 0.77 (0.55-1.09) |
| **OPERA** | EDSS 12-week | 1655 | 827 | 828 | 0.60 (0.45-0.81) |
| **OPERA** | EDSS 24-week | 1655 | 827 | 828 | 0.60 (0.43-0.84) |
| **ORATORIO** | EDSS 12-week | 732 | 488 | 244 | 0.76 (0.59-0.98) |
| **ORATORIO** | EDSS 24-week | 732 | 488 | 244 | 0.75 (0.58-0.98) |

*ADJUSTED OR; CDW: confirmed disability worsening; EDSS: Expanded Disability Status Scale; T25FWT: timed 25-foot walk test; 9HPT: 9-hole peg test; HR: hazard ratio; CI: confidence interval;

**Supplementary Table 2. Rate of disability worsening event according to single and composite endpoints**

|  |  |  | **12-week CDW** | | | | **24-week CDW** | | | |
| --- | --- | --- | --- | --- | --- | --- | --- | --- | --- | --- |
|  |  |  | **TREATMENT ARM** | | **PLACEBO ARM** | | **TREATMENT ARM** | | **PLACEBO ARM** | |
| **Trial** | **MSType** | **CDW components** | **Total number of patients** | **Number (%) of events** | **Total number of patients** | **Number (%) of events** | **Total number of patients** | **Number (%) of events** | **Total number of patients** | **Number (%) of events** |
| ASCEND | SPMS | EDSS | 440 | 104 (23.64) | 449 | 105 (23.39) | 440 | 77 (17.5) | 449 | 87 (19.38) |
| ASCEND | SPMS | 9HPT (mean hand) | 440 | 60 (13.64) | 449 | 89 (19.82) | 440 | 44 (10) | 449 | 61 (13.59) |
| ASCEND | SPMS | T25FW | 440 | 202 (45.91) | 449 | 201 (44.77) | 440 | 151 (34.32) | 449 | 150 (33.41) |
| ASCEND | SPMS | EDSS ∪ T25FW ∪ 9HPT | 440 | 241 (54.77) | 449 | 255 (56.79) | 440 | 191 (43.41) | 449 | 203 (45.21) |
| ASCEND | SPMS | EDSS ∪ T25FW | 440 | 236 (53.64) | 449 | 237 (52.78) | 440 | 184 (41.82) | 449 | 187 (41.65) |
| ASCEND | SPMS | EDSS ∪ 9HPT | 440 | 135 (30.68) | 449 | 156 (34.74) | 440 | 102 (23.18) | 449 | 126 (28.06) |
| ASCEND | SPMS | EDSS ∩ 9HPT | 440 | 29 (6.59) | 449 | 38 (8.46) | 440 | 19 (4.32) | 449 | 22 (4.9) |
| ASCEND | SPMS | EDSS ∩ T25FWT | 440 | 70 (15.91) | 449 | 69 (15.37) | 440 | 44 (10) | 449 | 50 (11.14) |
| ASCEND | SPMS | EDSS ∩ (T25FWT ∪ 9HPT) | 440 | 75 (17.05) | 449 | 79 (17.59) | 440 | 49 (11.14) | 449 | 59 (13.14) |
| ASCEND | SPMS | ≥2 of (EDSS, T25FWT, 9HPT) | 440 | 101 (22.95) | 449 | 112 (24.94) | 440 | 67 (15.23) | 449 | 82 (18.26) |
| ASCEND | SPMS | EDSS ∩ T25FWT ∩ 9HPT | 440 | 24 (5.45) | 449 | 28 (6.24) | 440 | 14 (3.18) | 449 | 13 (2.9) |
| ASCEND | SPMS | cognitive measure | 439 | 37 (8.43) | 449 | 46 (10.24) | 439 | 17 (3.87) | 449 | 27 (6.01) |
| ASCEND | SPMS | EDSS ∪ T25FWT ∪ 9HPT ∪ COG | 439 | 253 (57.63) | 449 | 270 (60.13) | 439 | 196 (44.65) | 449 | 214 (47.66) |
| ASCEND | SPMS | EDSS ∪ T25FWT ∪ COG | 439 | 248 (56.49) | 449 | 254 (56.57) | 439 | 189 (43.05) | 449 | 199 (44.32) |
| ASCEND | SPMS | EDSS ∪ 9HPT ∪ COG | 439 | 156 (35.54) | 449 | 180 (40.09) | 439 | 112 (25.51) | 449 | 142 (31.63) |
| ASCEND | SPMS | EDSS ∩ 9HPT ∩ COG | 439 | 3 (0.68) | 449 | 7 (1.56) | 439 | 0 (0) | 449 | 4 (0.89) |
| ASCEND | SPMS | EDSS ∩ T25FWT ∩ COG | 439 | 5 (1.14) | 449 | 9 (2) | 439 | 0 (0) | 449 | 4 (0.89) |
| ASCEND | SPMS | (EDSS ∩ T25FWT) ∪ (EDSS ∩ 9HPT) ∪ (EDSS ∩ COG) | 440 | 79 (17.95) | 449 | 80 (17.82) | 440 | 51 (11.59) | 449 | 60 (13.36) |
| ASCEND | SPMS | (EDSS ∩ T25FWT) ∪ (EDSS ∩ 9HPT) ∪ (T25FWT ∩ 9HPT) ∪ (EDSS ∩ COG) ∪ (T25FWT ∩ COG) ∪ (9HPT ∩ COG) | 440 | 12 (2.73) | 449 | 19 (4.23) | 440 | 5 (1.14) | 449 | 9 (2) |
| ASCEND | SPMS | EDSS ∩ T25FWT ∩ 9HPT ∩ COG | 439 | 3 (0.68) | 449 | 5 (1.11) | 439 | 0 (0) | 449 | 2 (0.45) |
| BRAVO | RRMS | EDSS | 434 | 41 (9.45) | 450 | 60 (13.33) | 434 | 28 (6.45) | 450 | 48 (10.67) |
| BRAVO | RRMS | 9HPT (mean hand) | 434 | 24 (5.53) | 449 | 27 (6.01) | 434 | 24 (5.53) | 449 | 27 (6.01) |
| BRAVO | RRMS | T25FW | 434 | 94 (21.66) | 450 | 90 (20) | 434 | 66 (15.21) | 450 | 63 (14) |
| BRAVO | RRMS | EDSS ∪ T25FW ∪ 9HPT | 434 | 126 (29.03) | 449 | 133 (29.62) | 434 | 94 (21.66) | 449 | 102 (22.72) |
| BRAVO | RRMS | EDSS ∪ T25FW | 434 | 114 (26.27) | 450 | 118 (26.22) | 434 | 82 (18.89) | 450 | 87 (19.33) |
| BRAVO | RRMS | EDSS ∪ 9HPT | 434 | 58 (13.36) | 449 | 79 (17.59) | 434 | 45 (10.37) | 449 | 68 (15.14) |
| BRAVO | RRMS | EDSS ∩ 9HPT | 434 | 7 (1.61) | 449 | 8 (1.78) | 434 | 7 (1.61) | 449 | 7 (1.56) |
| BRAVO | RRMS | EDSS ∩ T25FWT | 434 | 21 (4.84) | 450 | 32 (7.11) | 434 | 12 (2.76) | 450 | 24 (5.33) |
| BRAVO | RRMS | EDSS ∩ (T25FWT ∪ 9HPT) | 434 | 22 (5.07) | 450 | 32 (7.11) | 434 | 14 (3.23) | 450 | 26 (5.78) |
| BRAVO | RRMS | ≥2 of (EDSS, T25FWT, 9HPT) | 434 | 27 (6.22) | 450 | 36 (8) | 434 | 19 (4.38) | 450 | 31 (6.89) |
| BRAVO | RRMS | EDSS ∩ T25FWT ∩ 9HPT | 434 | 6 (1.38) | 449 | 8 (1.78) | 434 | 5 (1.15) | 449 | 5 (1.11) |
| BRAVO | RRMS | cognitive measure | 434 | 50 (11.52) | 450 | 66 (14.67) | 434 | 48 (11.06) | 450 | 65 (14.44) |
| BRAVO | RRMS | EDSS ∪ T25FWT ∪ 9HPT ∪ COG | 434 | 156 (35.94) | 450 | 171 (38) | 434 | 127 (29.26) | 450 | 143 (31.78) |
| BRAVO | RRMS | EDSS ∪ 9HPT ∪ COG | 434 | 97 (22.35) | 450 | 123 (27.33) | 434 | 85 (19.59) | 450 | 114 (25.33) |
| BRAVO | RRMS | EDSS ∪ T25FWT ∪ COG | 434 | 145 (33.41) | 450 | 161 (35.78) | 434 | 116 (26.73) | 450 | 133 (29.56) |
| BRAVO | RRMS | EDSS ∩ 9HPT ∩ COG | 434 | 1 (0.23) | 449 | 3 (0.67) | 434 | 1 (0.23) | 449 | 3 (0.67) |
| BRAVO | RRMS | EDSS ∩ T25FWT ∩ COG | 434 | 8 (1.84) | 450 | 11 (2.44) | 434 | 5 (1.15) | 450 | 9 (2) |
| BRAVO | RRMS | (EDSS ∩ T25FWT) ∪ (EDSS ∩ 9HPT) ∪ (EDSS ∩ COG) | 434 | 23 (5.3) | 450 | 37 (8.22) | 434 | 15 (3.46) | 450 | 30 (6.67) |
| BRAVO | RRMS | (EDSS ∩ T25FWT) ∪ (EDSS ∩ 9HPT) ∪ (T25FWT ∩ 9HPT) ∪ (EDSS ∩ COG) ∪ (T25FWT ∩ COG) ∪ (9HPT ∩ COG) | 434 | 9 (2.07) | 450 | 12 (2.67) | 434 | 6 (1.38) | 450 | 10 (2.22) |
| BRAVO | RRMS | EDSS ∩ T25FWT ∩ 9HPT ∩ COG | 434 | 1 (0.23) | 449 | 3 (0.67) | 434 | 1 (0.23) | 449 | 3 (0.67) |
| CONFIRM | RRMS | EDSS | 707 | 165 (23.34) | 363 | 86 (23.69) | 707 | 143 (20.23) | 363 | 73 (20.11) |
| CONFIRM | RRMS | 9HPT (mean hand) | 707 | 38 (5.37) | 363 | 27 (7.44) | 707 | 24 (3.39) | 363 | 17 (4.68) |
| CONFIRM | RRMS | T25FW | 707 | 119 (16.83) | 363 | 64 (17.63) | 707 | 81 (11.46) | 363 | 40 (11.02) |
| CONFIRM | RRMS | EDSS ∪ T25FW ∪ 9HPT | 707 | 254 (35.93) | 363 | 133 (36.64) | 707 | 200 (28.29) | 363 | 105 (28.93) |
| CONFIRM | RRMS | EDSS ∪ 9HPT | 707 | 184 (26.03) | 363 | 99 (27.27) | 707 | 156 (22.07) | 363 | 83 (22.87) |
| CONFIRM | RRMS | EDSS ∪ T25FW | 707 | 239 (33.8) | 363 | 122 (33.61) | 707 | 190 (26.87) | 363 | 96 (26.45) |
| CONFIRM | RRMS | EDSS ∩ 9HPT | 707 | 19 (2.69) | 363 | 14 (3.86) | 707 | 11 (1.56) | 363 | 7 (1.93) |
| CONFIRM | RRMS | EDSS ∩ T25FWT | 707 | 45 (6.36) | 363 | 28 (7.71) | 707 | 34 (4.81) | 363 | 17 (4.68) |
| CONFIRM | RRMS | EDSS ∩ (T25FWT ∪ 9HPT) | 707 | 54 (7.64) | 363 | 34 (9.37) | 707 | 40 (5.66) | 363 | 20 (5.51) |
| CONFIRM | RRMS | ≥2 of (EDSS, T25FWT, 9HPT) | 707 | 58 (8.2) | 363 | 36 (9.92) | 707 | 43 (6.08) | 363 | 21 (5.79) |
| CONFIRM | RRMS | EDSS ∩ T25FWT ∩ 9HPT | 707 | 10 (1.41) | 363 | 8 (2.2) | 707 | 5 (0.71) | 363 | 4 (1.1) |
| CONFIRM | RRMS | cognitive measure | 707 | 72 (10.18) | 363 | 35 (9.64) | 707 | 43 (6.08) | 363 | 24 (6.61) |
| CONFIRM | RRMS | EDSS ∪ T25FWT ∪ 9HPT ∪ COG | 707 | 292 (41.3) | 363 | 149 (41.05) | 707 | 223 (31.54) | 363 | 118 (32.51) |
| CONFIRM | RRMS | EDSS ∪ T25FWT ∪ COG | 707 | 280 (39.6) | 363 | 140 (38.57) | 707 | 214 (30.27) | 363 | 111 (30.58) |
| CONFIRM | RRMS | EDSS ∪ 9HPT ∪ COG | 707 | 230 (32.53) | 363 | 121 (33.33) | 707 | 184 (26.03) | 363 | 100 (27.55) |
| CONFIRM | RRMS | EDSS ∩ 9HPT ∩ COG | 707 | 5 (0.71) | 363 | 3 (0.83) | 707 | 3 (0.42) | 363 | 1 (0.28) |
| CONFIRM | RRMS | EDSS ∩ T25FWT ∩ COG | 707 | 3 (0.42) | 363 | 5 (1.38) | 707 | 1 (0.14) | 363 | 1 (0.28) |
| CONFIRM | RRMS | (EDSS ∩ T25FWT) ∪ (EDSS ∩ 9HPT) ∪ (EDSS ∩ COG) | 707 | 67 (9.48) | 363 | 38 (10.47) | 707 | 47 (6.65) | 363 | 23 (6.34) |
| CONFIRM | RRMS | (EDSS ∩ T25FWT) ∪ (EDSS ∩ 9HPT) ∪ (T25FWT ∩ 9HPT) ∪ (EDSS ∩ COG) ∪ (T25FWT ∩ COG) ∪ (9HPT ∩ COG) | 707 | 10 (1.41) | 363 | 7 (1.93) | 707 | 7 (0.99) | 363 | 2 (0.55) |
| CONFIRM | RRMS | EDSS ∩ T25FWT ∩ 9HPT ∩ COG | 707 | 1 (0.14) | 363 | 2 (0.55) | 707 | 0 (0) | 363 | 0 (0) |
| DEFINE | RRMS | EDSS | 826 | 202 (24.46) | 408 | 117 (28.68) | 826 | 176 (21.31) | 408 | 98 (24.02) |
| DEFINE | RRMS | 9HPT (mean hand) | 826 | 45 (5.45) | 408 | 22 (5.39) | 826 | 27 (3.27) | 408 | 9 (2.21) |
| DEFINE | RRMS | T25FW | 826 | 128 (15.5) | 408 | 65 (15.93) | 826 | 83 (10.05) | 408 | 42 (10.29) |
| DEFINE | RRMS | EDSS ∪ T25FW ∪ 9HPT | 826 | 299 (36.2) | 408 | 157 (38.48) | 826 | 242 (29.3) | 408 | 125 (30.64) |
| DEFINE | RRMS | EDSS ∪ 9HPT | 826 | 229 (27.72) | 408 | 129 (31.62) | 826 | 191 (23.12) | 408 | 104 (25.49) |
| DEFINE | RRMS | EDSS ∪ T25FW | 826 | 280 (33.9) | 408 | 150 (36.76) | 826 | 230 (27.85) | 408 | 121 (29.66) |
| DEFINE | RRMS | EDSS ∩ 9HPT | 826 | 18 (2.18) | 408 | 10 (2.45) | 826 | 12 (1.45) | 408 | 3 (0.74) |
| DEFINE | RRMS | EDSS ∩ T25FWT | 826 | 50 (6.05) | 408 | 32 (7.84) | 826 | 29 (3.51) | 408 | 19 (4.66) |
| DEFINE | RRMS | EDSS ∩ (T25FWT ∪ 9HPT) | 826 | 59 (7.14) | 408 | 35 (8.58) | 826 | 36 (4.36) | 408 | 20 (4.9) |
| DEFINE | RRMS | ≥2 of (EDSS, T25FWT, 9HPT) | 826 | 67 (8.11) | 408 | 40 (9.8) | 826 | 39 (4.72) | 408 | 22 (5.39) |
| DEFINE | RRMS | EDSS ∩ T25FWT ∩ 9HPT | 826 | 9 (1.09) | 408 | 7 (1.72) | 826 | 5 (0.61) | 408 | 2 (0.49) |
| DEFINE | RRMS | cognitive measure | 825 | 53 (6.42) | 407 | 25 (6.14) | 825 | 28 (3.39) | 407 | 14 (3.44) |
| DEFINE | RRMS | EDSS ∪ T25FWT ∪ 9HPT ∪ COG | 825 | 317 (38.42) | 407 | 166 (40.79) | 825 | 253 (30.67) | 407 | 131 (32.19) |
| DEFINE | RRMS | EDSS ∪ 9HPT ∪ COG | 825 | 255 (30.91) | 407 | 140 (34.4) | 825 | 208 (25.21) | 407 | 112 (27.52) |
| DEFINE | RRMS | EDSS ∪ T25FWT ∪ COG | 825 | 302 (36.61) | 407 | 159 (39.07) | 825 | 243 (29.45) | 407 | 127 (31.2) |
| DEFINE | RRMS | EDSS ∩ 9HPT ∩ COG | 825 | 6 (0.73) | 407 | 7 (1.72) | 825 | 2 (0.24) | 407 | 1 (0.25) |
| DEFINE | RRMS | EDSS ∩ T25FWT ∩ COG | 825 | 7 (0.85) | 407 | 5 (1.23) | 825 | 2 (0.24) | 407 | 3 (0.74) |
| DEFINE | RRMS | (EDSS ∩ T25FWT) ∪ (EDSS ∩ 9HPT) ∪ (EDSS ∩ COG) | 826 | 69 (8.35) | 408 | 42 (10.29) | 826 | 41 (4.96) | 408 | 23 (5.64) |
| DEFINE | RRMS | (EDSS ∩ T25FWT) ∪ (EDSS ∩ 9HPT) ∪ (T25FWT ∩ 9HPT) ∪ (EDSS ∩ COG) ∪ (T25FWT ∩ COG) ∪ (9HPT ∩ COG) | 826 | 13 (1.57) | 408 | 7 (1.72) | 826 | 4 (0.48) | 408 | 3 (0.74) |
| DEFINE | RRMS | EDSS ∩ T25FWT ∩ 9HPT ∩ COG | 825 | 3 (0.36) | 407 | 5 (1.23) | 825 | 1 (0.12) | 407 | 1 (0.25) |
| EXPAND | SPMS | EDSS | 1099 | 293 (26.66) | 546 | 180 (32.97) | 1099 | 223 (20.29) | 546 | 145 (26.56) |
| EXPAND | SPMS | 9HPT (mean hand) | 1096 | 168 (15.33) | 546 | 94 (17.22) | 1096 | 92 (8.39) | 546 | 56 (10.26) |
| EXPAND | SPMS | T25FW | 1096 | 420 (38.32) | 546 | 234 (42.86) | 1096 | 295 (26.92) | 546 | 170 (31.14) |
| EXPAND | SPMS | EDSS ∪ T25FW ∪ 9HPT | 1094 | 585 (53.47) | 546 | 322 (58.97) | 1094 | 445 (40.68) | 546 | 248 (45.42) |
| EXPAND | SPMS | EDSS ∪ 9HPT | 1096 | 398 (36.31) | 546 | 225 (41.21) | 1096 | 277 (25.27) | 546 | 172 (31.5) |
| EXPAND | SPMS | EDSS ∪ T25FW | 1097 | 537 (48.95) | 546 | 305 (55.86) | 1097 | 408 (37.19) | 546 | 234 (42.86) |
| EXPAND | SPMS | EDSS ∩ 9HPT | 1096 | 63 (5.75) | 546 | 49 (8.97) | 1096 | 38 (3.47) | 546 | 29 (5.31) |
| EXPAND | SPMS | EDSS ∩ T25FWT | 1096 | 176 (16.06) | 546 | 109 (19.96) | 1096 | 110 (10.04) | 546 | 81 (14.84) |
| EXPAND | SPMS | EDSS ∩ (T25FWT ∪ 9HPT) | 1099 | 197 (17.93) | 546 | 124 (22.71) | 1099 | 127 (11.56) | 546 | 94 (17.22) |
| EXPAND | SPMS | ≥2 of (EDSS, T25FWT, 9HPT) | 1099 | 254 (23.11) | 546 | 152 (27.84) | 1099 | 144 (13.1) | 546 | 107 (19.6) |
| EXPAND | SPMS | EDSS ∩ T25FWT ∩ 9HPT | 1093 | 42 (3.84) | 546 | 34 (6.23) | 1093 | 21 (1.92) | 546 | 16 (2.93) |
| EXPAND | SPMS | cognitive measure | 1096 | 224 (20.44) | 545 | 142 (26.06) | 1096 | 186 (16.97) | 545 | 119 (21.83) |
| EXPAND | SPMS | EDSS ∪ T25FWT ∪ 9HPT ∪ COG | 1093 | 675 (61.76) | 546 | 367 (67.22) | 1093 | 536 (49.04) | 546 | 297 (54.4) |
| EXPAND | SPMS | EDSS ∪ 9HPT ∪ COG | 1095 | 514 (46.94) | 546 | 302 (55.31) | 1094 | 394 (36.01) | 546 | 248 (45.42) |
| EXPAND | SPMS | EDSS ∪ T25FWT ∪ COG | 1095 | 640 (58.45) | 546 | 354 (64.84) | 1095 | 508 (46.39) | 546 | 286 (52.38) |
| EXPAND | SPMS | EDSS ∩ 9HPT ∩ COG | 1094 | 23 (2.1) | 545 | 15 (2.75) | 1094 | 14 (1.28) | 545 | 8 (1.47) |
| EXPAND | SPMS | EDSS ∩ T25FWT ∩ COG | 1093 | 52 (4.76) | 545 | 29 (5.32) | 1093 | 27 (2.47) | 545 | 18 (3.3) |
| EXPAND | SPMS | (EDSS ∩ T25FWT) ∪ (EDSS ∩ 9HPT) ∪ (EDSS ∩ COG) | 1099 | 215 (19.56) | 546 | 140 (25.64) | 1099 | 146 (13.28) | 546 | 107 (19.6) |
| EXPAND | SPMS | (EDSS ∩ T25FWT) ∪ (EDSS ∩ 9HPT) ∪ (T25FWT ∩ 9HPT) ∪ (EDSS ∩ COG) ∪ (T25FWT ∩ COG) ∪ (9HPT ∩ COG) | 1097 | 77 (7.02) | 545 | 45 (8.26) | 1098 | 41 (3.73) | 545 | 27 (4.95) |
| EXPAND | SPMS | EDSS ∩ T25FWT ∩ 9HPT ∩ COG | 1091 | 14 (1.28) | 545 | 11 (2.02) | 1091 | 7 (0.64) | 545 | 5 (0.92) |
| INFORMS | PPMS | EDSS | 336 | 158 (47.02) | 487 | 238 (48.87) | 336 | 133 (39.58) | 487 | 209 (42.92) |
| INFORMS | PPMS | 9HPT (mean hand) | 336 | 85 (25.3) | 487 | 127 (26.08) | 336 | 57 (16.96) | 487 | 97 (19.92) |
| INFORMS | PPMS | T25FW | 336 | 185 (55.06) | 487 | 265 (54.41) | 336 | 154 (45.83) | 487 | 228 (46.82) |
| INFORMS | PPMS | EDSS ∪ T25FW ∪ 9HPT | 336 | 233 (69.35) | 487 | 338 (69.4) | 336 | 201 (59.82) | 487 | 300 (61.6) |
| INFORMS | PPMS | EDSS ∪ 9HPT | 336 | 190 (56.55) | 487 | 272 (55.85) | 336 | 158 (47.02) | 487 | 239 (49.08) |
| INFORMS | PPMS | EDSS ∪ T25FW | 336 | 216 (64.29) | 487 | 321 (65.91) | 336 | 187 (55.65) | 487 | 285 (58.52) |
| INFORMS | PPMS | EDSS ∩ 9HPT | 336 | 53 (15.77) | 487 | 93 (19.1) | 336 | 32 (9.52) | 487 | 67 (13.76) |
| INFORMS | PPMS | EDSS ∩ T25FWT | 336 | 127 (37.8) | 487 | 182 (37.37) | 336 | 100 (29.76) | 487 | 152 (31.21) |
| INFORMS | PPMS | EDSS ∩ (T25FWT ∪ 9HPT) | 336 | 132 (39.29) | 487 | 196 (40.25) | 336 | 108 (32.14) | 487 | 163 (33.47) |
| INFORMS | PPMS | ≥2 of (EDSS, T25FWT, 9HPT) | 336 | 147 (43.75) | 487 | 213 (43.74) | 336 | 119 (35.42) | 487 | 178 (36.55) |
| INFORMS | PPMS | EDSS ∩ T25FWT ∩ 9HPT | 336 | 48 (14.29) | 487 | 79 (16.22) | 336 | 24 (7.14) | 487 | 56 (11.5) |
| INFORMS | PPMS | cognitive measure | 335 | 56 (16.72) | 487 | 66 (13.55) | 335 | 31 (9.25) | 487 | 38 (7.8) |
| INFORMS | PPMS | EDSS ∪ T25FWT ∪ 9HPT ∪ COG | 336 | 246 (73.21) | 487 | 347 (71.25) | 336 | 210 (62.5) | 487 | 304 (62.42) |
| INFORMS | PPMS | EDSS ∪ 9HPT ∪ COG | 336 | 207 (61.61) | 487 | 291 (59.75) | 336 | 168 (50) | 487 | 253 (51.95) |
| INFORMS | PPMS | EDSS ∪ T25FWT ∪ COG | 336 | 234 (69.64) | 487 | 331 (67.97) | 336 | 200 (59.52) | 487 | 290 (59.55) |
| INFORMS | PPMS | EDSS ∩ 9HPT ∩ COG | 335 | 15 (4.48) | 487 | 19 (3.9) | 335 | 6 (1.79) | 487 | 6 (1.23) |
| INFORMS | PPMS | EDSS ∩ T25FWT ∩ COG | 335 | 27 (8.06) | 487 | 29 (5.95) | 335 | 13 (3.88) | 487 | 16 (3.29) |
| INFORMS | PPMS | (EDSS ∩ T25FWT) ∪ (EDSS ∩ 9HPT) ∪ (EDSS ∩ COG) | 335 | 135 (40.3) | 487 | 202 (41.48) | 335 | 109 (32.54) | 487 | 166 (34.09) |
| INFORMS | PPMS | (EDSS ∩ T25FWT) ∪ (EDSS ∩ 9HPT) ∪ (T25FWT ∩ 9HPT) ∪ (EDSS ∩ COG) ∪ (T25FWT ∩ COG) ∪ (9HPT ∩ COG) | 336 | 31 (9.23) | 487 | 40 (8.21) | 336 | 16 (4.76) | 487 | 20 (4.11) |
| INFORMS | PPMS | EDSS ∩ T25FWT ∩ 9HPT ∩ COG | 335 | 14 (4.18) | 487 | 15 (3.08) | 335 | 4 (1.19) | 487 | 6 (1.23) |
| OLYMPUS | PPMS | EDSS | 292 | 99 (33.9) | 147 | 57 (38.78) | 292 | 82 (28.08) | 147 | 48 (32.65) |
| OLYMPUS | PPMS | 9HPT (mean hand) | 291 | 56 (19.24) | 147 | 31 (21.09) | 291 | 35 (12.03) | 147 | 20 (13.61) |
| OLYMPUS | PPMS | T25FW | 292 | 117 (40.07) | 147 | 72 (48.98) | 292 | 88 (30.14) | 147 | 51 (34.69) |
| OLYMPUS | PPMS | EDSS ∪ T25FW ∪ 9HPT | 291 | 173 (59.45) | 147 | 92 (62.59) | 291 | 145 (49.83) | 147 | 76 (51.7) |
| OLYMPUS | PPMS | EDSS ∪ 9HPT | 291 | 129 (44.33) | 147 | 71 (48.3) | 291 | 103 (35.4) | 147 | 57 (38.78) |
| OLYMPUS | PPMS | EDSS ∪ T25FW | 292 | 158 (54.11) | 147 | 88 (59.86) | 292 | 132 (45.21) | 147 | 73 (49.66) |
| OLYMPUS | PPMS | EDSS ∩ 9HPT | 291 | 26 (8.93) | 147 | 17 (11.56) | 291 | 14 (4.81) | 147 | 11 (7.48) |
| OLYMPUS | PPMS | EDSS ∩ T25FWT | 292 | 58 (19.86) | 147 | 41 (27.89) | 292 | 38 (13.01) | 147 | 26 (17.69) |
| OLYMPUS | PPMS | EDSS ∩ (T25FWT ∪ 9HPT) | 292 | 65 (22.26) | 147 | 44 (29.93) | 292 | 42 (14.38) | 147 | 30 (20.41) |
| OLYMPUS | PPMS | ≥2 of (EDSS, T25FWT, 9HPT) | 292 | 80 (27.4) | 147 | 54 (36.73) | 292 | 50 (17.12) | 147 | 36 (24.49) |
| OLYMPUS | PPMS | EDSS ∩ T25FWT ∩ 9HPT | 291 | 19 (6.53) | 147 | 14 (9.52) | 291 | 10 (3.44) | 147 | 7 (4.76) |
| OPERA | RRMS | EDSS | 825 | 75 (9.09) | 828 | 129 (15.58) | 825 | 54 (6.55) | 828 | 95 (11.47) |
| OPERA | RRMS | 9HPT (mean hand) | 825 | 32 (3.88) | 826 | 40 (4.84) | 825 | 17 (2.06) | 826 | 31 (3.75) |
| OPERA | RRMS | T25FW | 825 | 118 (14.3) | 826 | 147 (17.8) | 825 | 75 (9.09) | 826 | 96 (11.62) |
| OPERA | RRMS | EDSS ∪ T25FW ∪ 9HPT | 823 | 179 (21.75) | 826 | 249 (30.15) | 823 | 126 (15.31) | 826 | 179 (21.67) |
| OPERA | RRMS | EDSS ∪ 9HPT | 824 | 101 (12.26) | 826 | 157 (19.01) | 824 | 68 (8.25) | 826 | 118 (14.29) |
| OPERA | RRMS | EDSS ∪ T25FW | 824 | 167 (20.27) | 826 | 231 (27.97) | 824 | 115 (13.96) | 826 | 161 (19.49) |
| OPERA | RRMS | EDSS ∩ 9HPT | 824 | 6 (0.73) | 826 | 12 (1.45) | 824 | 3 (0.36) | 826 | 8 (0.97) |
| OPERA | RRMS | EDSS ∩ T25FWT | 824 | 26 (3.16) | 826 | 45 (5.45) | 824 | 14 (1.7) | 826 | 30 (3.63) |
| OPERA | RRMS | EDSS ∩ (T25FWT ∪ 9HPT) | 826 | 29 (3.51) | 828 | 50 (6.04) | 826 | 17 (2.06) | 828 | 34 (4.11) |
| OPERA | RRMS | ≥2 of (EDSS, T25FWT, 9HPT) | 826 | 43 (5.21) | 826 | 60 (7.26) | 826 | 20 (2.42) | 826 | 39 (4.72) |
| OPERA | RRMS | EDSS ∩ T25FWT ∩ 9HPT | 823 | 3 (0.36) | 826 | 7 (0.85) | 823 | 0 (0) | 826 | 4 (0.48) |
| OPERA | RRMS | cognitive measure | 815 | 102 (12.52) | 811 | 160 (19.73) | 815 | 68 (8.34) | 811 | 102 (12.58) |
| OPERA | RRMS | EDSS ∪ T25FWT ∪ 9HPT ∪ COG | 815 | 247 (30.31) | 811 | 334 (41.18) | 813 | 175 (21.53) | 810 | 249 (30.74) |
| OPERA | RRMS | EDSS ∪ 9HPT ∪ COG | 814 | 184 (22.6) | 810 | 270 (33.33) | 814 | 128 (15.72) | 810 | 197 (24.32) |
| OPERA | RRMS | EDSS ∪ T25FWT ∪ COG | 816 | 237 (29.04) | 811 | 324 (39.95) | 814 | 165 (20.27) | 810 | 235 (29.01) |
| OPERA | RRMS | EDSS ∩ 9HPT ∩ COG | 814 | 2 (0.25) | 810 | 7 (0.86) | 814 | 1 (0.12) | 810 | 2 (0.25) |
| OPERA | RRMS | EDSS ∩ T25FWT ∩ COG | 814 | 8 (0.98) | 810 | 18 (2.22) | 814 | 3 (0.37) | 810 | 6 (0.74) |
| OPERA | RRMS | (EDSS ∩ T25FWT) ∪ (EDSS ∩ 9HPT) ∪ (EDSS ∩ COG) | 825 | 35 (4.24) | 828 | 67 (8.09) | 825 | 20 (2.42) | 828 | 44 (5.31) |
| OPERA | RRMS | (EDSS ∩ T25FWT) ∪ (EDSS ∩ 9HPT) ∪ (T25FWT ∩ 9HPT) ∪ (EDSS ∩ COG) ∪ (T25FWT ∩ COG) ∪ (9HPT ∩ COG) | 825 | 11 (1.33) | 826 | 22 (2.66) | 825 | 4 (0.48) | 826 | 9 (1.09) |
| OPERA | RRMS | EDSS ∩ T25FWT ∩ 9HPT ∩ COG | 813 | 1 (0.12) | 810 | 5 (0.62) | 813 | 0 (0) | 810 | 1 (0.12) |
| ORATORIO | PPMS | EDSS | 488 | 188 (38.52) | 244 | 97 (39.75) | 488 | 160 (32.79) | 244 | 83 (34.02) |
| ORATORIO | PPMS | 9HPT (mean hand) | 488 | 89 (18.24) | 244 | 63 (25.82) | 488 | 70 (14.34) | 244 | 47 (19.26) |
| ORATORIO | PPMS | T25FW | 488 | 242 (49.59) | 244 | 129 (52.87) | 488 | 199 (40.78) | 244 | 105 (43.03) |
| ORATORIO | PPMS | EDSS ∪ T25FW ∪ 9HPT | 488 | 305 (62.5) | 244 | 159 (65.16) | 488 | 264 (54.1) | 244 | 142 (58.2) |
| ORATORIO | PPMS | EDSS ∪ 9HPT | 488 | 227 (46.52) | 244 | 121 (49.59) | 488 | 193 (39.55) | 244 | 103 (42.21) |
| ORATORIO | PPMS | EDSS ∪ T25FW | 488 | 285 (58.4) | 244 | 155 (63.52) | 488 | 246 (50.41) | 244 | 133 (54.51) |
| ORATORIO | PPMS | EDSS ∩ 9HPT | 488 | 50 (10.25) | 244 | 39 (15.98) | 488 | 37 (7.58) | 244 | 27 (11.07) |
| ORATORIO | PPMS | EDSS ∩ T25FWT | 488 | 145 (29.71) | 244 | 71 (29.1) | 488 | 113 (23.16) | 244 | 55 (22.54) |
| ORATORIO | PPMS | EDSS ∩ (T25FWT ∪ 9HPT) | 488 | 145 (29.71) | 244 | 74 (30.33) | 488 | 115 (23.57) | 244 | 60 (24.59) |
| ORATORIO | PPMS | ≥2 of (EDSS, T25FWT, 9HPT) | 488 | 164 (33.61) | 244 | 94 (38.52) | 488 | 130 (26.64) | 244 | 71 (29.1) |
| ORATORIO | PPMS | EDSS ∩ T25FWT ∩ 9HPT | 488 | 50 (10.25) | 244 | 36 (14.75) | 488 | 35 (7.17) | 244 | 22 (9.02) |
| ORATORIO | PPMS | cognitive measure | 488 | 89 (18.24) | 244 | 32 (13.11) | 488 | 52 (10.66) | 244 | 24 (9.84) |
| ORATORIO | PPMS | EDSS ∪ T25FWT ∪ 9HPT ∪ COG | 488 | 336 (68.85) | 244 | 170 (69.67) | 488 | 290 (59.43) | 244 | 153 (62.7) |
| ORATORIO | PPMS | EDSS ∪ 9HPT ∪ COG | 488 | 272 (55.74) | 244 | 136 (55.74) | 488 | 225 (46.11) | 244 | 118 (48.36) |
| ORATORIO | PPMS | EDSS ∪ T25FWT ∪ COG | 488 | 318 (65.16) | 244 | 167 (68.44) | 488 | 274 (56.15) | 244 | 146 (59.84) |
| ORATORIO | PPMS | EDSS ∩ 9HPT ∩ COG | 488 | 10 (2.05) | 244 | 10 (4.1) | 488 | 4 (0.82) | 244 | 4 (1.64) |
| ORATORIO | PPMS | EDSS ∩ T25FWT ∩ COG | 488 | 32 (6.56) | 244 | 10 (4.1) | 488 | 14 (2.87) | 244 | 3 (1.23) |
| ORATORIO | PPMS | (EDSS ∩ T25FWT) ∪ (EDSS ∩ 9HPT) ∪ (EDSS ∩ COG) | 488 | 153 (31.35) | 244 | 76 (31.15) | 488 | 118 (24.18) | 244 | 61 (25) |
| ORATORIO | PPMS | (EDSS ∩ T25FWT) ∪ (EDSS ∩ 9HPT) ∪ (T25FWT ∩ 9HPT) ∪ (EDSS ∩ COG) ∪ (T25FWT ∩ COG) ∪ (9HPT ∩ COG) | 488 | 34 (6.97) | 244 | 14 (5.74) | 488 | 15 (3.07) | 244 | 6 (2.46) |
| ORATORIO | PPMS | EDSS ∩ T25FWT ∩ 9HPT ∩ COG | 488 | 10 (2.05) | 244 | 9 (3.69) | 488 | 4 (0.82) | 244 | 2 (0.82) |

CDW: confirmed disability worsening; EDSS: Expanded Disability Status Scale; T25FWT: timed 25-foot walk test; 9HPT: 9-hole peg test; COG: cognitive decline

**Supplementary Table 3. Differences in Z-statistics for Confirmed Disability Worsening endpoints based on EDSS, 9HPT, T25FWT and composite endpoints at 12- and 24-week confirmation intervals** **using a common definition of confirmed disability worsening across trials (default “msprog” package parameters).**

|  |  | **12-week CDW** | **24-week CDW** |
| --- | --- | --- | --- |
| **Trial** | **CDW endpoint** | **DeltaZ** | **DeltaZ** |
| ASCEND | EDSS | 0 | 0 |
| ASCEND | 9HPT (mean hand) | -2.601 | -0.963 |
| ASCEND | T25FW | -0.005 | 0.787 |
| ASCEND | EDSS ∪ T25FW ∪ 9HPT | -0.41 | 0.223 |
| ASCEND | EDSS ∪ T25FW | -1.08 | 0.012 |
| ASCEND | EDSS ∪ 9HPT | -1.23 | -0.903 |
| ASCEND | EDSS ∩ T25FW | -0.073 | 0.061 |
| ASCEND | EDSS ∩ 9HPT | -1.3 | 0.259 |
| ASCEND | EDSS ∩ (T25FWT ∪ 9HPT) | -0.524 | -0.278 |
| ASCEND | ≥2 of (EDSS, T25FWT, 9HPT) | -0.979 | -0.566 |
| ASCEND | EDSS ∩ T25FWT ∩ 9HPT | -0.738 | 0.938 |
| ASCEND | cognitive decline | -1.017 | -0.678 |
| ASCEND | EDSS ∪ T25FWT ∪ 9HPT ∪ COG | -0.685 | -0.197 |
| ASCEND | EDSS ∪ T25FWT ∪ COG | -1.169 | -0.392 |
| ASCEND | EDSS ∪ 9HPT ∪ COG | -1.378 | -1.286 |
| ASCEND | EDSS ∩ T25FW ∩ COG | -1.083 | 0.296 |
| ASCEND | EDSS ∩ 9HPT ∩ COG | -1.237 | 0.926 |
| ASCEND | (EDSS ∩ T25FWT) ∪ (EDSS ∩ 9HPT) ∪ (EDSS ∩ COG) | -0.273 | -0.197 |
| ASCEND | (EDSS ∩ T25FWT) ∪ (EDSS ∩ 9HPT) ∪ (T25FWT ∩ 9HPT) ∪ (EDSS ∩ COG) ∪ (T25FWT ∩ COG) ∪ (9HPT ∩ COG) | -1.323 | -0.244 |
| ASCEND | EDSS ∩ T25FWT ∩ 9HPT ∩ COG | -0.715 | 0.926 |
| BRAVO | EDSS | 0 | 0 |
| BRAVO | 9HPT (mean hand) | 1.525 | 1.897 |
| BRAVO | T25FW | 2.476 | 2.686 |
| BRAVO | EDSS ∪ T25FW ∪ 9HPT | 1.767 | 1.912 |
| BRAVO | EDSS ∪ T25FW | 2.34 | 2.442 |
| BRAVO | EDSS ∪ 9HPT | 0.081 | 0.117 |
| BRAVO | EDSS ∩ T25FW | 0.374 | 0.266 |
| BRAVO | EDSS ∩ 9HPT | 1.617 | 2.244 |
| BRAVO | EDSS ∩ (T25FWT ∪ 9HPT) | 0.53 | 0.371 |
| BRAVO | ≥2 of (EDSS, T25FWT, 9HPT) | 0.759 | 0.568 |
| BRAVO | EDSS ∩ T25FWT ∩ 9HPT | 1.335 | 2.231 |
| BRAVO | cognitive decline | 0.406 | 0.66 |
| BRAVO | EDSS ∪ T25FWT ∪ 9HPT ∪ COG | 1.327 | 1.472 |
| BRAVO | EDSS ∪ T25FWT ∪ COG | 1.566 | 1.7 |
| BRAVO | EDSS ∪ 9HPT ∪ COG | 0.07 | 0.168 |
| BRAVO | EDSS ∩ T25FW ∩ COG | 1.183 | 1.17 |
| BRAVO | EDSS ∩ 9HPT ∩ COG | 0.884 | 1.256 |
| BRAVO | (EDSS ∩ T25FWT) ∪ (EDSS ∩ 9HPT) ∪ (EDSS ∩ COG) | 0.081 | 0.04 |
| BRAVO | (EDSS ∩ T25FWT) ∪ (EDSS ∩ 9HPT) ∪ (T25FWT ∩ 9HPT) ∪ (EDSS ∩ COG) ∪ (T25FWT ∩ COG) ∪ (9HPT ∩ COG) | 1.22 | 1.241 |
| BRAVO | EDSS ∩ T25FWT ∩ 9HPT ∩ COG | 0.884 | 1.255 |
| CONFIRM | EDSS | 0 | 0 |
| CONFIRM | 9HPT (mean hand) | -0.918 | -0.641 |
| CONFIRM | T25FW | 0.084 | 0.653 |
| CONFIRM | EDSS ∪ T25FW ∪ 9HPT | 0.193 | 0.085 |
| CONFIRM | EDSS ∪ T25FW | -0.314 | 0.22 |
| CONFIRM | EDSS ∪ 9HPT | -0.139 | -0.081 |
| CONFIRM | EDSS ∩ T25FW | -0.591 | 0.296 |
| CONFIRM | EDSS ∩ 9HPT | -0.761 | -0.192 |
| CONFIRM | EDSS ∩ (T25FWT ∪ 9HPT) | -0.727 | 0.298 |
| CONFIRM | ≥2 of (EDSS, T25FWT, 9HPT) | -0.669 | 0.415 |
| CONFIRM | EDSS ∩ T25FWT ∩ 9HPT | -0.663 | -0.395 |
| CONFIRM | cognitive decline | 0.752 | 0.074 |
| CONFIRM | EDSS ∪ T25FWT ∪ 9HPT ∪ COG | 0.675 | 0.053 |
| CONFIRM | EDSS ∪ T25FWT ∪ COG | 0.467 | 0.24 |
| CONFIRM | EDSS ∪ 9HPT ∪ COG | 0.211 | -0.241 |
| CONFIRM | EDSS ∩ T25FW ∩ COG | -1.322 | -0.194 |
| CONFIRM | EDSS ∩ 9HPT ∩ COG | 0.044 | 0.599 |
| CONFIRM | (EDSS ∩ T25FWT) ∪ (EDSS ∩ 9HPT) ∪ (EDSS ∩ COG) | -0.293 | 0.364 |
| CONFIRM | (EDSS ∩ T25FWT) ∪ (EDSS ∩ 9HPT) ∪ (T25FWT ∩ 9HPT) ∪ (EDSS ∩ COG) ∪ (T25FWT ∩ COG) ∪ (9HPT ∩ COG) | -0.322 | 0.995 |
| CONFIRM | EDSS ∩ T25FWT ∩ 9HPT ∩ COG | -0.859 |  |
| DEFINE | EDSS | 0 | 0 |
| DEFINE | 9HPT (mean hand) | 2.504 | 2.981 |
| DEFINE | T25FW | 2.385 | 1.901 |
| DEFINE | EDSS ∪ T25FW ∪ 9HPT | 1.82 | 1.545 |
| DEFINE | EDSS ∪ T25FW | 2.83 | 2.447 |
| DEFINE | EDSS ∪ 9HPT | 0.908 | 0.997 |
| DEFINE | EDSS ∩ T25FW | 0.984 | 0.814 |
| DEFINE | EDSS ∩ 9HPT | 1.954 | 2.885 |
| DEFINE | EDSS ∩ (T25FWT ∪ 9HPT) | 1.219 | 1.321 |
| DEFINE | ≥2 of (EDSS, T25FWT, 9HPT) | 1.142 | 1.257 |
| DEFINE | EDSS ∩ T25FWT ∩ 9HPT | 1.449 | 2.122 |
| DEFINE | cognitive decline | 2.652 | 1.928 |
| DEFINE | EDSS ∪ T25FWT ∪ 9HPT ∪ COG | 1.856 | 1.473 |
| DEFINE | EDSS ∪ T25FWT ∪ COG | 2.647 | 2.309 |
| DEFINE | EDSS ∪ 9HPT ∪ COG | 1.184 | 1.043 |
| DEFINE | EDSS ∩ T25FW ∩ COG | 1.672 | 0.696 |
| DEFINE | EDSS ∩ 9HPT ∩ COG | 0.8 | 1.891 |
| DEFINE | (EDSS ∩ T25FWT) ∪ (EDSS ∩ 9HPT) ∪ (EDSS ∩ COG) | 0.953 | 1.246 |
| DEFINE | (EDSS ∩ T25FWT) ∪ (EDSS ∩ 9HPT) ∪ (T25FWT ∩ 9HPT) ∪ (EDSS ∩ COG) ∪ (T25FWT ∩ COG) ∪ (9HPT ∩ COG) | 2.179 | 1.365 |
| DEFINE | EDSS ∩ T25FWT ∩ 9HPT ∩ COG | 0.663 | 1.398 |
| EXPAND | EDSS | 0 | 0 |
| EXPAND | 9HPT (mean hand) | 1.849 | 1.777 |
| EXPAND | T25FW | 0.781 | 0.959 |
| EXPAND | EDSS ∪ T25FW ∪ 9HPT | 0.273 | 0.762 |
| EXPAND | EDSS ∪ T25FW | 0.899 | 1.068 |
| EXPAND | EDSS ∪ 9HPT | 0.648 | 0.17 |
| EXPAND | EDSS ∩ T25FW | 0.69 | 0.071 |
| EXPAND | EDSS ∩ 9HPT | 0.355 | 1.207 |
| EXPAND | EDSS ∩ (T25FWT ∪ 9HPT) | 0.316 | -0.238 |
| EXPAND | ≥2 of (EDSS, T25FWT, 9HPT) | 0.485 | -0.55 |
| EXPAND | EDSS ∩ T25FWT ∩ 9HPT | 0.631 | 1.669 |
| EXPAND | cognitive decline | 0.18 | 0.518 |
| EXPAND | EDSS ∪ T25FWT ∪ 9HPT ∪ COG | -0.096 | 0.317 |
| EXPAND | EDSS ∪ T25FWT ∪ COG | 0.149 | 0.505 |
| EXPAND | EDSS ∪ 9HPT ∪ COG | -0.594 | -0.853 |
| EXPAND | EDSS ∩ T25FW ∩ COG | 2.247 | 1.988 |
| EXPAND | EDSS ∩ 9HPT ∩ COG | 1.957 | 2.671 |
| EXPAND | (EDSS ∩ T25FWT) ∪ (EDSS ∩ 9HPT) ∪ (EDSS ∩ COG) | -0.218 | -0.428 |
| EXPAND | (EDSS ∩ T25FWT) ∪ (EDSS ∩ 9HPT) ∪ (T25FWT ∩ 9HPT) ∪ (EDSS ∩ COG) ∪ (T25FWT ∩ COG) ∪ (9HPT ∩ COG) | 1.823 | 1.789 |
| EXPAND | EDSS ∩ T25FWT ∩ 9HPT ∩ COG | 1.641 | 2.355 |
| INFORMS | EDSS | 0 | 0 |
| INFORMS | 9HPT (mean hand) | 0.532 | -0.119 |
| INFORMS | T25FW | 0.745 | 0.674 |
| INFORMS | EDSS ∪ T25FW ∪ 9HPT | 0.398 | 0.475 |
| INFORMS | EDSS ∪ T25FW | 0.826 | 0.996 |
| INFORMS | EDSS ∪ 9HPT | 0.599 | 0.338 |
| INFORMS | EDSS ∩ T25FW | 0.649 | 0.432 |
| INFORMS | EDSS ∩ 9HPT | -0.402 | -0.886 |
| INFORMS | EDSS ∩ (T25FWT ∪ 9HPT) | 0.387 | 0.535 |
| INFORMS | ≥2 of (EDSS, T25FWT, 9HPT) | 0.707 | 0.637 |
| INFORMS | EDSS ∩ T25FWT ∩ 9HPT | 0.025 | -1.144 |
| INFORMS | cognitive decline | 1.834 | 1.413 |
| INFORMS | EDSS ∪ T25FWT ∪ 9HPT ∪ COG | 0.955 | 0.762 |
| INFORMS | EDSS ∪ T25FWT ∪ COG | 1.375 | 1.268 |
| INFORMS | EDSS ∪ 9HPT ∪ COG | 0.958 | 0.235 |
| INFORMS | EDSS ∩ T25FW ∩ COG | 1.702 | 1.244 |
| INFORMS | EDSS ∩ 9HPT ∩ COG | 1.006 | 1.449 |
| INFORMS | (EDSS ∩ T25FWT) ∪ (EDSS ∩ 9HPT) ∪ (EDSS ∩ COG) | 0.389 | 0.471 |
| INFORMS | (EDSS ∩ T25FWT) ∪ (EDSS ∩ 9HPT) ∪ (T25FWT ∩ 9HPT) ∪ (EDSS ∩ COG) ∪ (T25FWT ∩ COG) ∪ (9HPT ∩ COG) | 1.061 | 1.208 |
| INFORMS | EDSS ∩ T25FWT ∩ 9HPT ∩ COG | 1.403 | 0.756 |
| OLYMPUS | EDSS | 0 | 0 |
| OLYMPUS | 9HPT (mean hand) | 0.451 | 0.414 |
| OLYMPUS | T25FW | -0.83 | -0.318 |
| OLYMPUS | EDSS ∪ T25FW ∪ 9HPT | 0.175 | 0.321 |
| OLYMPUS | EDSS ∪ T25FW | -0.221 | 0.024 |
| OLYMPUS | EDSS ∪ 9HPT | -0.02 | 0.171 |
| OLYMPUS | EDSS ∩ T25FW | -0.944 | -0.454 |
| OLYMPUS | EDSS ∩ 9HPT | 0.136 | -0.194 |
| OLYMPUS | EDSS ∩ (T25FWT ∪ 9HPT) | -0.778 | -0.697 |
| OLYMPUS | ≥2 of (EDSS, T25FWT, 9HPT) | -1.147 | -0.972 |
| OLYMPUS | EDSS ∩ T25FWT ∩ 9HPT | -0.103 | 0.243 |
| OPERA | EDSS | 0 | 0 |
| OPERA | 9HPT (mean hand) | 3.049 | 1.563 |
| OPERA | T25FW | 1.72 | 1.652 |
| OPERA | EDSS ∪ T25FW ∪ 9HPT | -0.168 | 0.045 |
| OPERA | EDSS ∪ T25FW | 1.283 | 1.178 |
| OPERA | EDSS ∪ 9HPT | 0.212 | -0.338 |
| OPERA | EDSS ∩ T25FW | -1.442 | -1.614 |
| OPERA | EDSS ∩ 9HPT | -0.474 | -0.603 |
| OPERA | EDSS ∩ (T25FWT ∪ 9HPT) | 1.632 | 1.181 |
| OPERA | ≥2 of (EDSS, T25FWT, 9HPT) | 2.256 | 1.067 |
| OPERA | EDSS ∩ T25FWT ∩ 9HPT | 2.913 | 3.718 |
| OPERA | cognitive decline | -0.064 | 0.645 |
| OPERA | EDSS ∪ T25FWT ∪ 9HPT ∪ COG | -0.889 | -0.861 |
| OPERA | EDSS ∪ T25FWT ∪ COG | -0.099 | -0.254 |
| OPERA | EDSS ∪ 9HPT ∪ COG | -0.87 | -0.822 |
| OPERA | EDSS ∩ T25FW ∩ COG | -1.029 | -0.134 |
| OPERA | EDSS ∩ 9HPT ∩ COG | -0.609 | 0.319 |
| OPERA | (EDSS ∩ T25FWT) ∪ (EDSS ∩ 9HPT) ∪ (EDSS ∩ COG) | 0.823 | 0.568 |
| OPERA | (EDSS ∩ T25FWT) ∪ (EDSS ∩ 9HPT) ∪ (T25FWT ∩ 9HPT) ∪ (EDSS ∩ COG) ∪ (T25FWT ∩ COG) ∪ (9HPT ∩ COG) | 2.19 | 2.276 |
| OPERA | EDSS ∩ T25FWT ∩ 9HPT ∩ COG | 2.707 | 3.719 |
| ORATORIO | EDSS | 0 | 0 |
| ORATORIO | 9HPT (mean hand) | -2.051 | -1.377 |
| ORATORIO | T25FW | -0.577 | -0.33 |
| ORATORIO | EDSS ∪ T25FW ∪ 9HPT | -0.112 | -0.555 |
| ORATORIO | EDSS ∪ T25FW | -0.272 | -0.723 |
| ORATORIO | EDSS ∪ 9HPT | -0.261 | -0.285 |
| ORATORIO | EDSS ∩ T25FW | 0.395 | 0.509 |
| ORATORIO | EDSS ∩ 9HPT | -1.951 | -1.189 |
| ORATORIO | EDSS ∩ (T25FWT ∪ 9HPT) | 0.045 | -0.019 |
| ORATORIO | ≥2 of (EDSS, T25FWT, 9HPT) | -0.982 | -0.411 |
| ORATORIO | EDSS ∩ T25FWT ∩ 9HPT | -1.52 | -0.487 |
| ORATORIO | cognitive decline | 2.237 | 0.898 |
| ORATORIO | EDSS ∪ T25FWT ∪ 9HPT ∪ COG | 0.586 | -0.114 |
| ORATORIO | EDSS ∪ T25FWT ∪ COG | 0.539 | -0.234 |
| ORATORIO | EDSS ∪ 9HPT ∪ COG | 0.831 | 0.068 |
| ORATORIO | EDSS ∩ T25FW ∩ COG | 1.684 | 1.852 |
| ORATORIO | EDSS ∩ 9HPT ∩ COG | -1.181 | -0.433 |
| ORATORIO | (EDSS ∩ T25FWT) ∪ (EDSS ∩ 9HPT) ∪ (EDSS ∩ COG) | 0.209 | 0.034 |
| ORATORIO | (EDSS ∩ T25FWT) ∪ (EDSS ∩ 9HPT) ∪ (T25FWT ∩ 9HPT) ∪ (EDSS ∩ COG) ∪ (T25FWT ∩ COG) ∪ (9HPT ∩ COG) | 0.985 | 0.969 |
| ORATORIO | EDSS ∩ T25FWT ∩ 9HPT ∩ COG | -0.913 | 0.551 |

z: Wald z-statistic, used to test whether the log(HR) differs significantly from zero CDW: confirmed disability worsening; EDSS: Expanded Disability Status Scale; T25FWT: timed 25-foot walk test; 9HPT: 9-hole peg test; COG: cognitive decline

**Supplementary Table 4. Differences in treatment effect Z-values (ΔZ relative to EDSS) and interaction p-values comparing exploratory composite disability worsening endpoints with EDSS alone in the overall population, RRMS, and progressive MS.**

|  |  | **12-week CDW** | | **24-week CDW** | |
| --- | --- | --- | --- | --- | --- |
|  | **population** | **ΔZ** | **p x interaction** | **ΔZ** | **p x interaction** |
| EDSS | Overall | 0 | - | 0 | - |
| T25FWT | Overall | 1.881 | 0.037 | 2.257 | 0.023 |
| 9HPT | Overall | 0.250 | 0.934 | 0.295 | 0.790 |
| EDSS ∪ T25FWT ∪ 9HPT | Overall | 1.555 | 0.012 | 1.757 | 0.004 |
| EDSS ∪ T25FWT | Overall | 1.369 | 0.020 | 1.683 | 0.004 |
| EDSS ∪ 9HPT | Overall | 0.594 | 0.140 | 0.350 | 0.331 |
| EDSS ∩ 9HPT | Overall | -1.516 | 0.052 | -0.402 | 0.221 |
| EDSS ∩ T25FWT | Overall | 0.446 | 0.577 | 0.193 | 0.880 |
| EDSS ∩ (T25FWT ∪ 9HPT) | Overall | 0.034 | 0.996 | -0.038 | 0.658 |
| ≥2 of (EDSS, T25FWT, 9HPT) | Overall | 0.065 | 0.930 | -0.263 | 0.548 |
| EDSS ∩ T25FWT ∩ 9HPT | Overall | -1.026 | 0.136 | 0.256 | 0.790 |
| EDSS | RRMS | 0 | - | 0 | - |
| T25FWT | RRMS | 2.269 | 0.016 | 2.176 | 0.029 |
| 9HPT | RRMS | 1.525 | 0.336 | 1.267 | 0.550 |
| EDSS ∪ T25FWT ∪ 9HPT | RRMS | 1.572 | 0.002 | 1.350 | 0.004 |
| EDSS ∪ T25FWT | RRMS | 1.607 | 0.001 | 1.419 | 0.001 |
| EDSS ∪ 9HPT | RRMS | 0.583 | 0.077 | 0.390 | 0.185 |
| EDSS ∩ 9HPT | RRMS | 0.380 | 0.857 | 1.710 | 0.497 |
| EDSS ∩ T25FWT | RRMS | -0.331 | 0.430 | -0.198 | 0.332 |
| EDSS ∩ (T25FWT ∪ 9HPT) | RRMS | -0.140 | 0.593 | 0.202 | 0.666 |
| ≥2 of (EDSS, T25FWT, 9HPT) | RRMS | 0.492 | 0.840 | 0.328 | 0.811 |
| EDSS ∩ T25FWT ∩ 9HPT | RRMS | -0.174 | 0.489 | 1.742 | 0.835 |
| EDSS | PROG | 0 | - | 0 | - |
| T25FWT | PROG | 0.522 | 0.562 | 1.210 | 0.280 |
| 9HPT | PROG | -0.938 | 0.325 | -0.750 | 0.396 |
| EDSS ∪ T25FWT ∪ 9HPT | PROG | 0.602 | 0.378 | 1.150 | 0.143 |
| EDSS ∪ T25FWT | PROG | 0.274 | 0.614 | 0.920 | 0.205 |
| EDSS ∪ 9HPT | PROG | 0.237 | 0.604 | 0.030 | 0.804 |
| EDSS ∩ 9HPT | PROG | -2.365 | 0.010 | -1.650 | 0.051 |
| EDSS ∩ T25FWT | PROG | 0.322 | 0.687 | 0.030 | 0.898 |
| EDSS ∩ (T25FWT ∪ 9HPT) | PROG | -0.274 | 0.605 | -0.550 | 0.394 |
| ≥2 of (EDSS, T25FWT, 9HPT) | PROG | -0.600 | 0.435 | -0.980 | 0.272 |
| EDSS ∩ T25FWT ∩ 9HPT | PROG | -1.602 | 0.069 | -0.700 | 0.190 |

CDW: confirmed disability worsening; EDSS: Expanded Disability Status Scale; T25FWT: timed 25-foot walk test; 9HPT: 9-hole peg test; RRMS: relapsing-remitting multiple sclerosis; PROG: progressive multiple sclerosis;

**Supplementary Table 5. Differences in treatment effect Z-values (ΔZ relative to EDSS) and interaction p-values comparing exploratory composite disability worsening endpoints including cognitive assessment with EDSS alone in the overall population, RRMS, and progressive MS.**

|  |  | **12-week CDW** | | **24-week CDW** | |
| --- | --- | --- | --- | --- | --- |
|  | **population** | **ΔZ** | **p x interaction** | **ΔZ** | **p x interaction** |
| EDSS | Overall | 0 | - | 0 | - |
| EDSS ∪ T25FWT ∪ 9HPT ∪ COG | Overall | 1.47 | 0.023 | 1.623 | 0.031 |
| EDSS ∪ T25FWT ∪ COG | Overall | 1.419 | 0.028 | 1.594 | 0.030 |
| EDSS ∪ 9HPT ∪ COG | Overall | 4.978 | 0.324 | 0.372 | 0.888 |
| EDSS ∩ 9HPT ∩ COG | Overall | -0.171 | 0.188 | 2.634 | 0.764 |
| EDSS ∩ T25FWT ∩ COG | Overall | 1.879 | 0.535 | 2.22 | 0.959 |
| (EDSS ∩ T25FWT) ∪ (EDSS ∩ 9HPT) ∪ (EDSS ∩ COG) | Overall | -0.082 | 0.914 | -0.177 | 0.609 |
| (EDSS ∩ T25FWT) ∪ (EDSS ∩ 9HPT) ∪ (T25FWT ∩ 9HPT) ∪ (EDSS ∩ COG) ∪ (T25FWT ∩ COG) ∪ (9HPT ∩ COG) | Overall | 1.249 | 0.830 | 1.723 | 0.947 |
| EDSS ∩ T25FWT ∩ 9HPT ∩ COG | Overall | -0.056 | 0.200 | 2.086 | 0.479 |
| EDSS | RRMS | 0 | - | 0 | - |
| EDSS ∪ T25FWT ∪ 9HPT ∪ COG | RRMS | 1.539 | 0.011 | 1.371 | 0.049 |
| EDSS ∪ T25FWT ∪ COG | RRMS | 1.489 | 0.013 | 1.332 | 0.049 |
| EDSS ∪ 9HPT ∪ COG | RRMS | 0.861 | 0.196 | 0.696 | 0.390 |
| EDSS ∩ 9HPT ∩ COG | RRMS | -1.64 | 0.130 | 1.603 | 0.899 |
| EDSS ∩ T25FWT ∩ COG | RRMS | -1.002 | 0.155 | -0.064 | 0.256 |
| (EDSS ∩ T25FWT) ∪ (EDSS ∩ 9HPT) ∪ (EDSS ∩ COG) | RRMS | -0.413 | 0.318 | -0.157 | 0.385 |
| (EDSS ∩ T25FWT) ∪ (EDSS ∩ 9HPT) ∪ (T25FWT ∩ 9HPT) ∪ (EDSS ∩ COG) ∪ (T25FWT ∩ COG) ∪ (9HPT ∩ COG) | RRMS | 0.188 | 0.597 | 0.911 | 0.744 |
| EDSS ∩ T25FWT ∩ 9HPT ∩ COG | RRMS | -2.981 | 0.031 | 0.437 | 0.311 |
| EDSS | PROG | 0 | - | 0 | - |
| EDSS ∪ T25FWT ∪ 9HPT ∪ COG | PROG | 0.733 | 0.345 | 1.039 | 0.223 |
| EDSS ∪ T25FWT ∪ COG | PROG | 0.707 | 0.356 | 1.04 | 0.213 |
| EDSS ∪ 9HPT ∪ COG | PROG | 0.288 | 0.728 | -0.197 | 0.674 |
| EDSS ∩ 9HPT ∩ COG | PROG | -0.124 | 0.411 | 2.036 | 0.721 |
| EDSS ∩ T25FWT ∩ COG | PROG | 2.193 | 0.181 | 2.599 | 0.623 |
| (EDSS ∩ T25FWT) ∪ (EDSS ∩ 9HPT) ∪ (EDSS ∩ COG) | PROG | -0.106 | 0.814 | -0.352 | 0.529 |
| (EDSS ∩ T25FWT) ∪ (EDSS ∩ 9HPT) ∪ (T25FWT ∩ 9HPT) ∪ (EDSS ∩ COG) ∪ (T25FWT ∩ COG) ∪ (9HPT ∩ COG) | PROG | 1.017 | 0.776 | 1.717 | 0.903 |
| EDSS ∩ T25FWT ∩ 9HPT ∩ COG | PROG | 0.183 | 0.610 | 1.73 | 0.716 |

CDW: confirmed disability worsening; EDSS: Expanded Disability Status Scale; T25FWT: timed 25-foot walk test; 9HPT: 9-hole peg test; RRMS: relapsing-remitting multiple sclerosis; PROG: progressive multiple sclerosis;
