## Supplementary material for "Composite endpoints to detect treatment effects on MS disability progression. Lessons from phase III trial data": Tables

**Table 1. Rate of 24-week confirmed disability worsening event according to single and composite endpoints**

|  | | | **TREATMENT ARM** | | **PLACEBO ARM** | |
| --- | --- | --- | --- | --- | --- | --- |
| **Trial** | **MS Type** | **CDW components** | **Total number of patients** | **Number (%) of events** | **Total number of patients** | **Number (%) of events** |
| ASCEND | SPMS | EDSS | 440 | 77 (17.5) | 449 | 87 (19.38) |
|  |  | T25FWT |  | 151 (34.32) |  | 150 (33.41) |
|  |  | 9HPT (mean hand) |  | 44 (10) |  | 61 (13.59) |
|  |  | EDSS ∪ T25FW ∪ 9HPT |  | 191 (43.41) |  | 203 (45.21) |
|  |  | EDSS ∪ T25FW |  | 163 (37.05) |  | 175 (38.98) |
|  |  | EDSS ∪ 9HPT |  | 102 (23.18) |  | 126 (28.06) |
| BRAVO | RRMS | EDSS | 434 | 28 (6.45) | 450 | 48 (10.67) |
|  |  | T25FWT |  | 66 (15.21) |  | 63 (14) |
|  |  | 9HPT (mean hand) |  | 24 (5.53) |  | 27 (6.01) |
|  |  | EDSS ∪ T25FW ∪ 9HPT |  | 94 (21.66) |  | 102 (22.72) |
|  |  | EDSS ∪ T25FW |  | 80 (18.43) |  | 80 (17.82) |
|  |  | EDSS ∪ 9HPT |  | 45 (10.37) |  | 68 (15.14) |
| CONFIRM | RRMS | EDSS | 707 | 143 (20.23) | 363 | 73 (20.11) |
|  |  | T25FWT |  | 81 (11.46) |  | 40 (11.02) |
|  |  | 9HPT (mean hand) |  | 24 (3.39) |  | 17 (4.68) |
|  |  | EDSS ∪ T25FW ∪ 9HPT |  | 200 (28.29) |  | 105 (28.93) |
|  |  | EDSS ∪ T25FW |  | 97 (13.72) |  | 52 (14.33) |
|  |  | EDSS ∪ 9HPT |  | 156 (22.07) |  | 83 (22.87) |
| DEFINE | RRMS | EDSS | 826 | 176 (21.31) | 408 | 98 (24.02) |
|  |  | T25FWT |  | 83 (10.05) |  | 42 (10.29) |
|  |  | 9HPT (mean hand) |  | 27 (3.27) |  | 9 (2.21) |
|  |  | EDSS ∪ T25FW ∪ 9HPT |  | 242 (29.3) |  | 125 (30.64) |
|  |  | EDSS ∪ T25FW |  | 102 (12.35) |  | 47 (11.52) |
|  |  | EDSS ∪ 9HPT |  | 191 (23.12) |  | 104 (25.49) |
| EXPAND | SPMS | EDSS | 1099 | 223 (20.29) | 546 | 145 (26.56) |
|  |  | T25FWT |  | 295 (26.92) |  | 170 (31.14) |
|  |  | 9HPT (mean hand) |  | 92 (8.39) |  | 56 (10.26) |
|  |  | EDSS ∪ T25FW ∪ 9HPT |  | 445 (40.68) |  | 248 (45.42) |
|  |  | EDSS ∪ T25FW |  | 349 (31.9) |  | 197 (36.08) |
|  |  | EDSS ∪ 9HPT |  | 277 (25.27) |  | 172 (31.5) |
| INFORMS | PPMS | EDSS | 336 | 133 (39.58) | 487 | 209 (42.92) |
|  |  | T25FWT |  | 154 (45.83) |  | 228 (46.82) |
|  |  | 9HPT (mean hand) |  | 57 (16.96) |  | 97 (19.92) |
|  |  | EDSS ∪ T25FW ∪ 9HPT |  | 201 (59.82) |  | 300 (61.6) |
|  |  | EDSS ∪ T25FW |  | 176 (52.38) |  | 254 (52.16) |
|  |  | EDSS ∪ 9HPT |  | 158 (47.02) |  | 239 (49.08) |
| OLYMPUS | PPMS | EDSS | 292 | 82 (28.08) | 147 | 48 (32.65) |
|  |  | T25FWT |  | 88 (30.14) |  | 51 (34.69) |
|  |  | 9HPT (mean hand) |  | 35 (12.03) |  | 20 (13.61) |
|  |  | EDSS ∪ T25FW ∪ 9HPT |  | 145 (49.83) |  | 76 (51.7) |
|  |  | EDSS ∪ T25FW |  | 105 (36.08) |  | 58 (39.46) |
|  |  | EDSS ∪ 9HPT |  | 103 (35.4) |  | 57 (38.78) |
| OPERA | RRMS | EDSS | 825 | 54 (6.55) | 828 | 95 (11.47) |
|  |  | T25FWT |  | 75 (9.09) |  | 96 (11.62) |
|  |  | 9HPT (mean hand) |  | 17 (2.06) |  | 31 (3.75) |
|  |  | EDSS ∪ T25FW ∪ 9HPT |  | 126 (15.31) |  | 179 (21.67) |
|  |  | EDSS ∪ T25FW |  | 89 (10.8) |  | 118 (14.29) |
|  |  | EDSS ∪ 9HPT |  | 68 (8.25) |  | 118 (14.29) |
| ORATORIO | PPMS | EDSS | 488 | 160 (32.79) | 244 | 83 (34.02) |
|  |  | T25FWT |  | 199 (40.78) |  | 105 (43.03) |
|  |  | 9HPT (mean hand) |  | 70 (14.34) |  | 47 (19.26) |
|  |  | EDSS ∪ T25FW ∪ 9HPT |  | 264 (54.1) |  | 142 (58.2) |
|  |  | EDSS ∪ T25FW |  | 219 (44.88) |  | 119 (48.77) |
|  |  | EDSS ∪ 9HPT |  | 193 (39.55) |  | 103 (42.21) |

CDW: confirmed disability worsening; EDSS: Expanded Disability Status Scale; T25FWT: timed 25-foot walk test; 9HPT: 9-hole peg test; RRMS: relapsing-remitting multiple sclerosis; SPMS: secondary progressive multiple sclerosis; PPMS: primary progressive multiple sclerosis;

**Table 2. Component-level contribution to composite 24-week confirmed disability worsening endpoints, based on earliest time-to-event attribution.**

| **CDW endpoint** | **No. of patients** | **No. Of events** | **% EDSS-driven** | **% T25FWT-driven** | **% 9HPT-driven** |
| --- | --- | --- | --- | --- | --- |
| EDSS | 9,369 | 1,962 | 100 | - | - |
| T25FWT | 9,364 | 2,137 | - | 100 | - |
| 9HPT (mean hand) | 9,362 | 755 | - | - | 100 |
| EDSS ∪ T25FW ∪ 9HPT | 9,358 | 3,388 | 38.3 | 49.6 | 12.1 |
| EDSS ∪ T25FW | 9,359 | 3,151 | 43.5 | 56.5 | - |
| EDSS ∪ 9HPT | 9,361 | 2,363 | 76.6 | - | 23.4 |

CDW: confirmed disability worsening; EDSS: Expanded Disability Status Scale; T25FWT: timed 25-foot walk test; 9HPT: 9-hole peg test;

**Table 3. Differences in Z-statistics for confirmed disability worsening endpoints based on EDSS, 9HPT, T25FWT and composite endpoints with a 24-week confirmation.**

| **Trial** | **CDW endpoint** | **DeltaZ** |
| --- | --- | --- |
| ASCEND | EDSS | 0 |
|  | **9HPT (mean hand)** | **-0.963** |
|  | T25FW | 0.787 |
|  | EDSS ∪ T25FW ∪ 9HPT | 0.271 |
|  | EDSS ∪ T25FW | 0.012 |
|  | EDSS ∪ 9HPT | -0.903 |
| BRAVO | **EDSS** | **0** |
|  | 9HPT (mean hand) | 1.897 |
|  | T25FW | 2.686 |
|  | EDSS ∪ T25FW ∪ 9HPT | 1.912 |
|  | EDSS ∪ T25FW | 2.442 |
|  | EDSS ∪ 9HPT | 0.117 |
| CONFIRM | EDSS | 0 |
|  | **9HPT (mean hand)** | **-0.641** |
|  | T25FW | 0.653 |
|  | EDSS ∪ T25FW ∪ 9HPT | 0.085 |
|  | EDSS ∪ T25FW | 0.220 |
|  | EDSS ∪ 9HPT | -0.081 |
| DEFINE | **EDSS** | **0** |
|  | 9HPT (mean hand) | 2.98 |
|  | T25FW | 1.901 |
|  | EDSS ∪ T25FW ∪ 9HPT | 1.545 |
|  | EDSS ∪ T25FW | 2.447 |
|  | EDSS ∪ 9HPT | 0.997 |
| EXPAND | **EDSS** | **0** |
|  | 9HPT (mean hand) | 1.777 |
|  | T25FW | 0.959 |
|  | EDSS ∪ T25FW ∪ 9HPT | 0.762 |
|  | EDSS ∪ T25FW | 1.068 |
|  | EDSS ∪ 9HPT | 0.170 |
| INFORMS | EDSS | 0 |
|  | **9HPT (mean hand)** | **-0.119** |
|  | T25FW | 0.674 |
|  | EDSS ∪ T25FW ∪ 9HPT | 0.475 |
|  | EDSS ∪ T25FW | 0.996 |
|  | EDSS ∪ 9HPT | 0.339 |
| OLYMPUS | EDSS | 0 |
|  | 9HPT (mean hand) | 0.414 |
|  | **T25FW** | **-0.318** |
|  | EDSS ∪ T25FW ∪ 9HPT | 0.321 |
|  | EDSS ∪ T25FW | 0.024 |
|  | EDSS ∪ 9HPT | 0.171 |
| OPERA | EDSS | 0 |
|  | 9HPT (mean hand) | 1.563 |
|  | T25FW | 1.652 |
|  | EDSS ∪ T25FW ∪ 9HPT | 0.045 |
|  | EDSS ∪ T25FW | 1.178 |
|  | **EDSS ∪ 9HPT** | **-0.338** |
| ORATORIO | EDSS | 0 |
|  | **9HPT (mean hand)** | **-1.377** |
|  | T25FW | -0.330 |
|  | EDSS ∪ T25FW ∪ 9HPT | 0.055 |
|  | EDSS ∪ T25FW | -0.723 |
|  | EDSS ∪ 9HPT | -0.285 |

CDW: confirmed disability worsening; EDSS: Expanded Disability Status Scale; T25FWT: timed 25-foot walk test; 9HPT: 9-hole peg test;
